# DEVELOPMENT AND VALIDATION OF A MODEL FOR THE PREDICTION OF MORTALITY IN CHILDREN UNDER FIVE YEARS WITH CLINICAL PNEUMONIA IN RURAL GAMBIA

**DOI:** 10.1101/2021.08.04.21260737

**Authors:** Alexander Jarde, David J. Jeffries, Grant A. Mackenzie

**Affiliations:** Department of Statistics and Bioinformatics, Medical Research Council Unit The Gambia at the London School of Hygiene & Tropical Medicine; Fajara; The Gambia; Disease Control and Elimination, Medical Research Council Unit The Gambia at the London School of Hygiene & Tropical Medicine; Fajara; The Gambia

**Keywords:** pneumococcal disease, pneumococcal infections, pneumonia, prediction, prognosis, mortality, severity

## Abstract

**Background:** Pneumonia is the leading cause of death in children aged 1-59 months. Prediction models for child pneumonia mortality have been developed using regression methods but their performance is insufficient for clinical use.

**Methods:** We used a variety of machine learning methods to develop a predictive model for mortality in children with clinical pneumonia enrolled in population-based surveillance in the Basse Health and Demographic Surveillance System in rural Gambia (n=11,012). Four machine learning algorithms (support vector machine, random forest, artifical neural network, and regularized logistic regression) were implemented, fitting all possible combinations of two or more of 16 selected features. Models were shortlisted based on their training set performance, the number of included features, and the reliability of feature measurement. The final model was selected considering its clinical interpretability.

**Results:** When we applied the final model to the test set (55 deaths), the area under the Receiver Operating Characteristic Curve was 0.88 (95% confidence interval: 0.84, 0.91), sensitivity was 0.78 and specificity was 0.77.

**Conclusions:** Our evaluation of multiple machine learning methods combined with minimal and pragmatic feature selection led to a predictive model with very good performance. We plan further validation of our model in different populations.

---

Pneumonia accounted for an estimated 921,000 childhood deaths in 2015, more than any other single condition. ^1^ The clinical status of children with pneumonia can rapidly deteriorate but existing mortality prediction tools have limited reliability. ^2^ The challenges of pneumonia diagnosis, assessment of severity, and the prediction of mortality are most acutely felt in low-resource settings, where expert personnel and well-equipped facilities are lacking.

In high-income settings, several clinical prediction scores of pneumonia severity have been developed and proven useful to triage patients for higher levels of clinical care, both in adults ^3–5^ and, less often in children. ^6,7^ Due to the different characteristics of the population and health care services, these scales are poorly suited to low-income settings, ^6^ where the ability to predict mortality in children with clinical pneumonia would be valuable both in primary care, to ensure appropriate referral for admission, and also in secondary and tertiary care, for referral to more intensive clinical support. In this context, a number of models to predict mortality in children with clinical pneumonia have been developed, providing moderately good levels of prediction. ^2^ Most of these models were developed using regression methods such as logistic regression (e.g. the RISC ^8^ and PERCH ^2^ scores. While a strength of regression models is their interpretability, superiror classification accuracy can be obtained from machine learning algorithms such as random forest (RF), support vector machines (SVM), and artificial neural networks (ANN).^9^

Our goal in this study was to develop and validate a predictive machine learning model for pneumonia mortality in children using variables readily available to clinicians.

## METHODS

### Study population

We used population-based surveillance data from the Pneumococcal Surveillance Project from June 4^th^ 2009 until December 31st 2017.^10^ This project was conducted in the Basse Health & Demographic Surveillance System (BHDSS) in the rural east of The Gambia, and was established to document the impact of pneumococcal conjugate vaccination on the incidence of invasive pneumococcal disease and radiological pneumonia in children aged 2 months and older. It included patients resident in the Basse Health & Demographic Surveillance System who met surveillance criteria for suspected pneumonia, sepsis or meningitis and were referred to clinicians for standardized review. In addition, our dataset included patients from January 1st 2011 until December 31st 2017 who did not meet surveillance criteria for suspected pneumonia, sepsis or meningitis or were not resident in the Basse Health & Demographic Surveillance System but had standardized clinician review,effectively including all children admitted at the Basse Health Centre. This health centre is a secondary care facility, and the only major health centre in Upper River Region, serving the largely rural population of approximately 260,000 on both banks of the River Gambia. ^10^

Inclusion criteria were age 2-59 months and having clinical pneumonia, defined as cough or difficulty breathing accompanied by one or more of: raised respiratory rate for age (≥50 breaths/minute if less than one year old; ≥40 breaths/minute if older than one year), lower chest wall indrawing, inability to sit or feed, convulsions, or oxygen saturation <92 percent.

We split our dataset by time (temporal validation) into a training set containing 2/3 of the sample (from June 4^th^ 2009 to July 26^th^ 2015), and a test set with the remaining 1/3 of the sample (from July 27^th^ 2015 to December 31^st^ 2017). Splitting the data in this manner allowed us to maintain the chronology of the data. Performance was not sensitive to the training and testing temporal cut-off.

### Outcome and predictors

Our outcome of interest was hospital based mortality. Potential explanatory variables in the predictive modelling process based on their clinical relevance are summarised in Table 1 Haemoglobin concentration was excluded due to missing data. Nasal flaring and grunting were not included due to concerns of substantial subjectivity and inter-observer disagreement. In addition, ‘abnormal conscious state’ was discarded for being a near zero-variance predictor. ^11^ All variables were collected at the time of admission. Impossible or implausible values were removed and only subjects with data for all features were included in the analyses (complete-case analyses), which represented over 94 percent of the original dataset (Table 1, Figure 1).

**Table 1.**
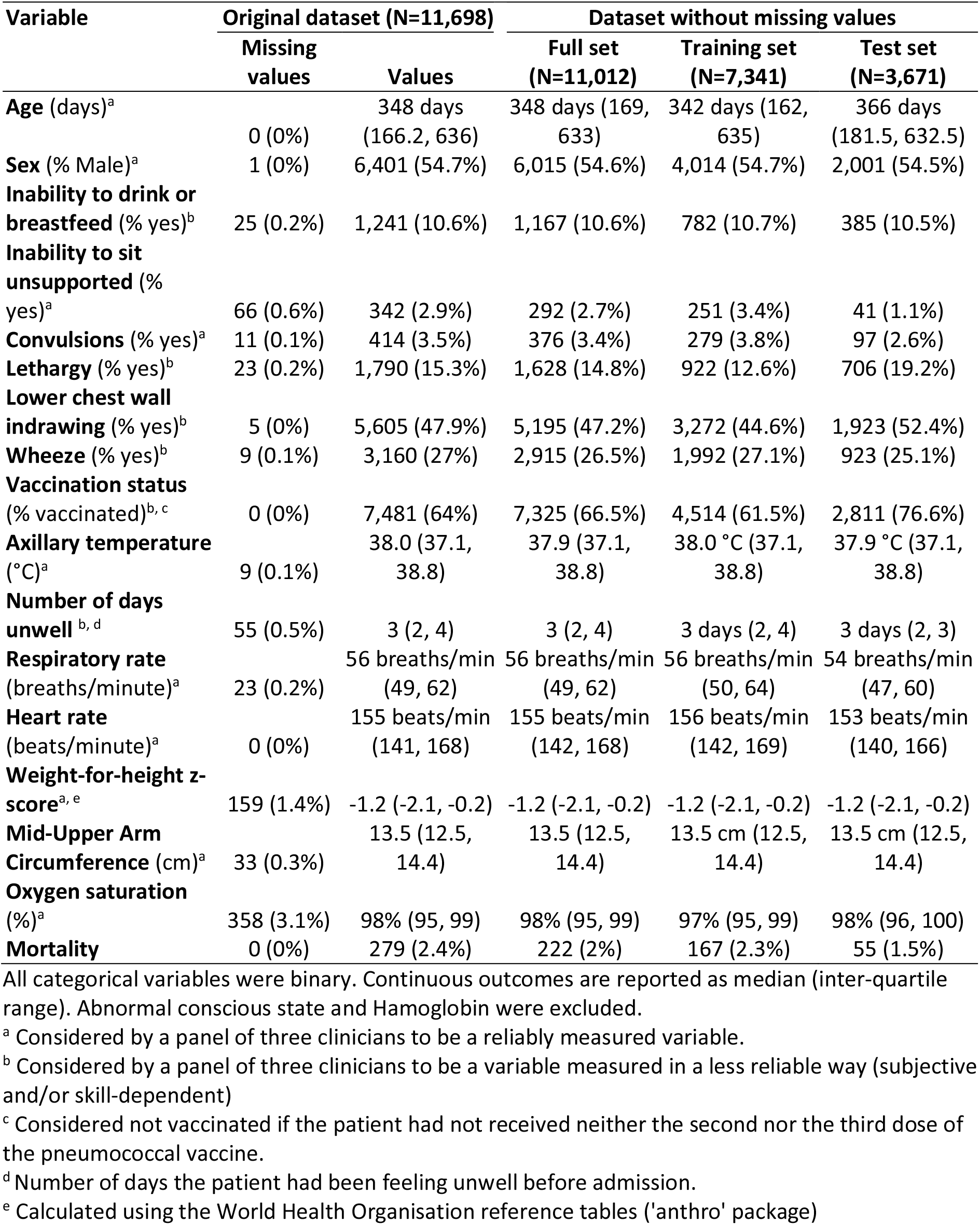
Distribution of Each Variable in the Original Dataset (Under-5 Children Admitted at the Basse Health Centre in Rural Gambia From 4th June2009 Until 31st December 2017) and the Subset Without Missing Values Used for the Development (Training Set) of Each Model and the Validation (Test Set) of the Final Model.

**Figure 1:**
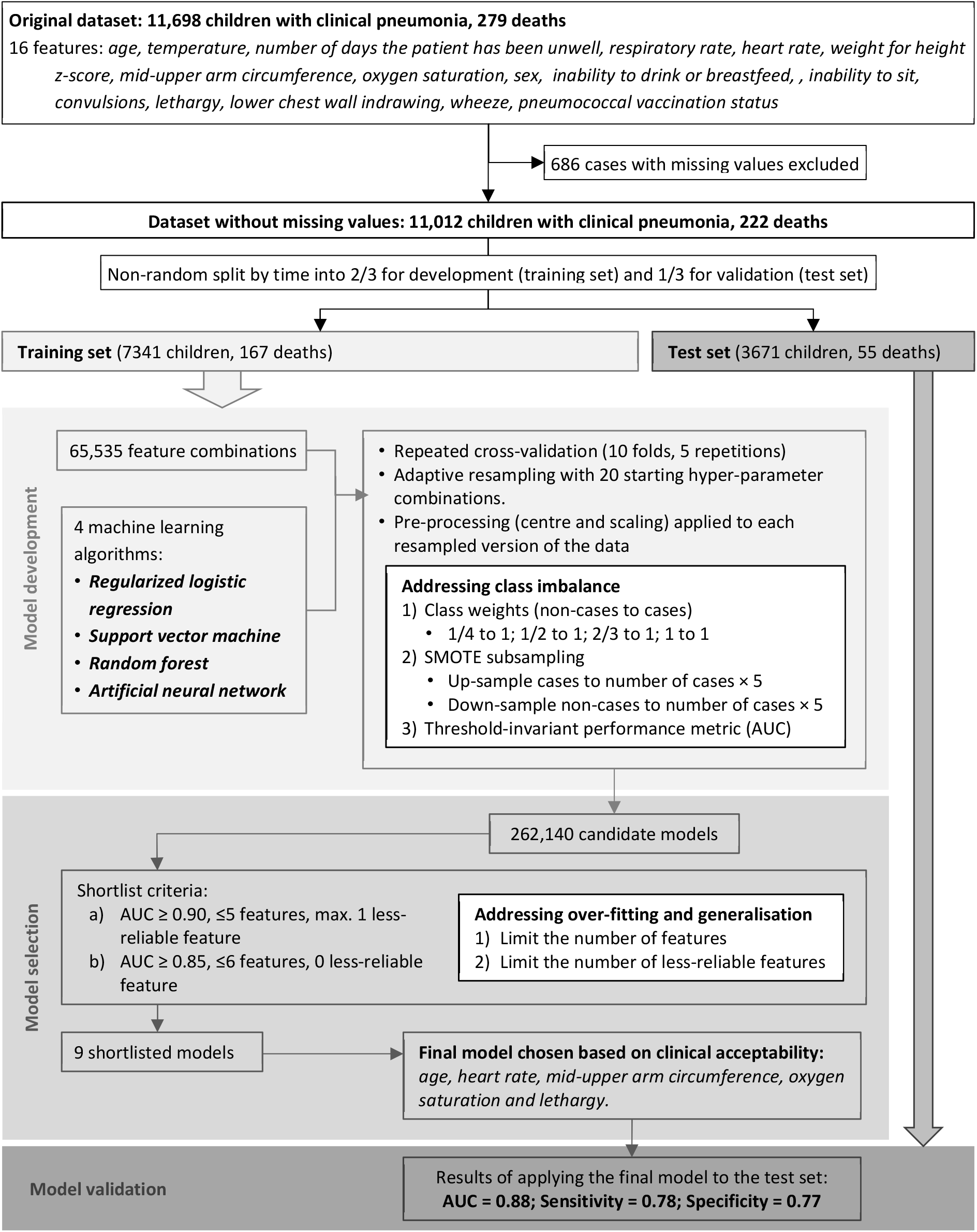
Study flow chart. Footnote: **SMOTE:** Synthetic Minority Over-Sampling Technique; **AUC:** Area Under the Receiver operating characteristic Curve

For this project, we asked a panel of three clinicians (one of the authors and Drs. Yekini-Ajao Olatunji and Galega Babila Lobga) to classify each feature based on reliability, the amount of skill required by the person measuring it, and consistency, with the agreed classification shown in Table 1.

### Model development

#### Types of Model

There is a large variety of machine learning algorithms, but, *a priori*, no best choice.^9^ Each has a distinct approach to the classification task: Random forests consist of multiple random decision trees, where each tree is built on a random sample from the original data and, at each tree node, a subset of features are randomly selected to generate the split that minimizes the probability that a randomly selected sample from a node will be incorrectly classified. ^12^ Support Vector Machines aim to find a boundary that separates two groups in the data by maximising the sum of the distance between the boundary and the chosen support vectors. ^13^ Artificial Neural Networks consist of an input layer, an output layer and a set of hidden layers with a certain number of neurons. Neurons in each layer are connected by sets of weights. The algorithm’s goal is to find the combination of weights that will minimize the amount of errors in the output layer. ^14^ Finally, we also used Regularized Logistic Regression (RLR), which uses maximum-likelihood estimation to find the coefficient values that minimize the error in the predicted probabilities. As part of the regression family, this method is most similar to the algorithm used during the development of previous scales, while allowing us to penalise increasing numbers of features (to limit overfitting) and apply different weights to the misclassification of cases and non-cases. ^15^

#### Feature selection during modelling

Since the number of potential predictors in our dataset was relatively low (16), we applied each of the selected machine learning algorithms to every possible combination of two or more features, resulting in a total of 65,519 feature combinations. We therefore did not perform further predictor selection. Given our exhaustive approach to the feature selection, we did not include interaction terms to avoid the risk of overfitting. ^16^

We explored the presence of non-linear relationships with fractional polynomials, but saw no consistent improvement in fit. Consequently, all continuous predictors were fitted as linear terms (centred and scaled by subtracting the mean and dividing by the standard deviation).

#### Sample size

Since this was a post-hoc analysis, no power calculation was possible. A rule of thumb to have at least 10 outcome events per model feature has been suggested by some authors based on empirical research. ^17–19^ With over 200 events and a maximum of 16 parameters, our dataset meets this criterion.

#### Model generation and internal validation

Using the training set, we generated four models for each of the 65,519 feature combinations (one for each of the machine learning algorithms considered). Each model was developed using repeated 10-fold cross-validation (5 repetitions) with adaptive resampling to optimize hyper-parameter tuning(starting with a random set of 20 hyper-parameter combinations each time). ^20,21^ The data set is highly imbalanced (only 2 percent of patients died) and we took the following precautions to avoid bias towards the majority class (survivors): First, for each algorithm four different weighing schemes penalised misclassification of death (four times more, two times more, 1.5 times more, or without penalty). Second, we used the Synthetic Minority Over-Sampling Technique ^22^ to balance the number of events and non-events by up-sampling the deaths and under-sumpling the number of non-events (survivors). To avoid overfitting with minority over-sampling, we limited the number of up-sampling of deaths to five times the original number of cases, forcing the down-sampling of non-event cases to result in an equal number of events (after up-sampling) and non-events. Third, we used a threshold-invariant metric -the Area Under the receiver operating characteristic Curve (AUC)-to avoid depending on threshold values when assessing the models’ performance on the training set (Figure 1).

For each model, we also identified the best threshold to classify patients (in the training set) by identifying the point in the receiver operating characteristic curve closest to the top-left corner (perfect sensitivity and specificity). All models were generated using R software, and the caret package. ^23,24^ Due to the high number of feature combinations, we ran our code in an embarassingly parallel format on a high performance computer.

#### Final model selectio

In order to reduce the risk of over-fitting and to increase the generalisability of our final model we did not rely solely on the performance metrics of the generated models during training, but also on the number of features and the number of less-reliable features (Table 1) included in the model. We therefore shortlisted, from the 262,076 candidate models (65,519 feature combinations in each of the four machine learning algorithms), those that either a) had an AUC ≥ 0.90, five or less features, and none or only one less-reliable feature; or b) had an AUC ≥ 0.85, six or less features, and no less-reliable feature. Initially, we chose the final model based on the the clinicalrelevance of features. Despite this model’s excellent performance when we validated it on the test set (see details in Supplementary file), the effect of some variables on the predicted probabilities was counter-intuitive (i.e. increasing temperature values resulted in decreasing predicted probabilities of death when all other variables were the same). We therefore chose another shortlisted model without counter-intuitive trends, thus improving the model’s interpretability. Further details can be found in the eAppendix.

#### Final model validation

Once the final model was chosen we evaluated its performance in the held-out test set: we used the model to generate for each subject a prediction probability of mortality, using the best threshold from the training set to classify them as dead or alive, and compared these predictions with the actual outcomes. We generated measures of calibration (density and calibration plots), discrimination (AUC) and classification (sensitivity and specificity). Calibration evaluates the agreement between a model’s prediction and the true outcome. Discrimination is the model’s ability to differentiate between survivors and deaths. Classification refers to the performance of the model after introducing a probability threshold. ^16^ We also generated the model’s learning curve, which consists of two curves generated by assessing the performance (AUC in our case) of the model when applied to the training set and the test set, respectively, at increasing fractions of the training set. The shape and dynamics of a learning curve can be used to assess if the model was overfitting (lines getting closer but then separating again over increased fractions of the training set used) and how well it was generalising (distance between the two lines). ^25^

## RESULTS

The initial dataset consisted of 11,698 subjects, of which 686 (5.9%) were excluded due to missing values (Figure 1). The remaining 11,012 subjects were non-randomly split into a training set (7,341 children, 167 deaths) and a test set (3,671 children, 55 deaths) based on their date of admission. Table 1 describes the characteristics of the participants before and after removing missing values, as well as in the training and test sets separately.

Among the 7,341 children in the training set, the median age was 342 days (Inter-Quartile Range 162-635), 54.7 percent were male, the median duration of hospitalisation was 2 days (Inter-Quartile Range 0 to 3), and 167 (2.3 percent) died in hospital.

The algorithm that resulted in most models with an AUC ≥ 0.9 was RF (3,527), followed by SVM (2,650), NNET (730), and RLR (131), with the number of features ranging from five to 16 and all including at least one feature considered less reliable. Only six of these models (2 RF, 2 SVM and 2 NNET) were shortlisted (5 or less features, with a maximum of one being considered less reliable. Relaxing the AUC criterion slightly, 60,048 models had an AUC ≥0.85 (RF 17,600, NNET 14,074, SVM 14905, RLR 13,469), with 10 of them having none of the less-reliable features. Limiting the total number of features to six or less resulted in shortlisting three more models (1 SVM, 1 NNET, 1 RF).

Consequently, only nine of the 262,140 candidate models were shortlisted based on their performance, their number of features and the reliability of measurement of features. Of these, the final model (a RF model which had an AUC=0.90, with sensitivity = 0.83 and specificity = 0.83) was chosen based on the included features and theirinterpretability: age, heart rate, mid-upper arm circumference, oxygen saturation and lethargy.

When we validated our final model on the test set, the AUC was 0.88 (95% confidence interval: 0.84 to 0.91). Figure 2A) shows the density of the real outcomes according to the model’s predicted probability of death and Figure 2B) shows the incidence of deaths among subjects in each decile of predicted probability. When we classified subjects in the test set applying the threshold probability 0.44 (the best threshold value identified during the development of the model using the training set), the resulting sensitivity and specificity were 0.78 and 0.77, respectively (Table 2).

**Figure 2:**
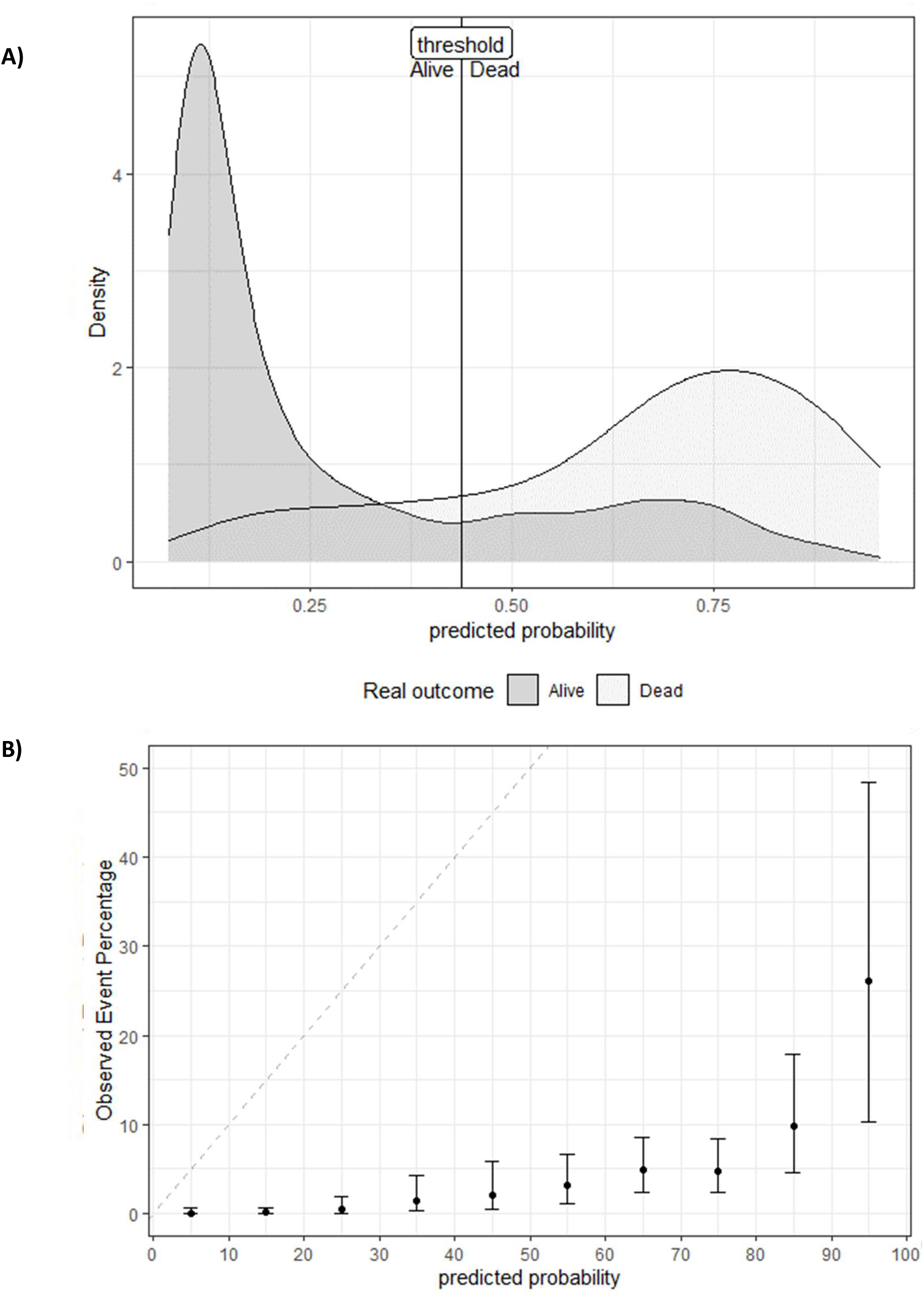
Density plot for real outcomes based on the predicted event probability estimates from the final model (panel A) and calibration plot (panel B).

**Table 2.** Confusion Matrix When Applying the Final Model to the Test Set For its Validation, Using the Threshold Established During the Development Phase Using the Training Set.

| **Predicted**<br>**outcomes** | **Real outcomes (column %)** |  |
| --- | --- | --- |
|  | **Alive** | **Dead** |
| **Alive** | 2,795 (77.3%) | 12 (21.8%) |
| **Dead** | 821 (22.7%) | 43 (78.2%) |

The learning curve (Figure 3) shows the evolution of the AUC when the final model was fitted using increasingly bigger fractions of the training set and then applied to that fraction of the training set and the complete test set. The curve indicates that the model reached stable performance (further training would not have substantially improved the model), with a relatively small and stable gap between the training and testing curve, indicating little overfitting and good generalisation.

**Figure 3:**
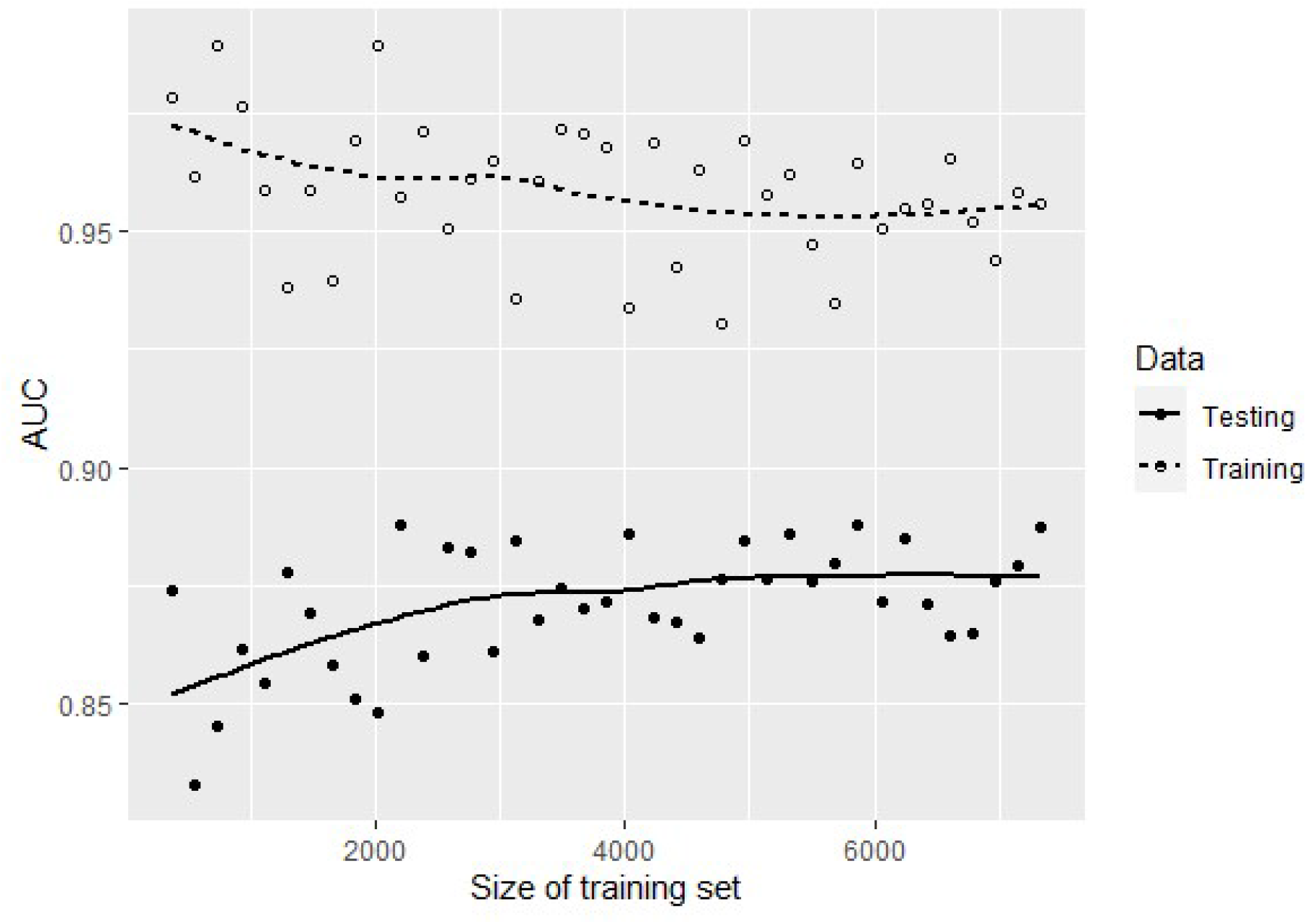
Learning curve of the model depicting the performance of the model with increasing size of training sets (circles and dotted line) and the held-out test set (full circles and continuous line).

The model implemented via a Shiny web can be downloaded from the supplemental digital content (as well as github: https://github.com/MRCG-djeffries/mortality-prediction) both as an independent .rds file or as a Shiny App, which incorporates a user interface that allows for individual and batch mortality predictions, as well as validation of our model in new datasets. Users can input values manually or upload a dataset to obtain the predicted probability of death for each record, as well as a dichotomous classification based on the optimal threshold of 0.44 identified in our training set. Finally, if data on mortality are provided in the dataset, it also allows assessment of model performance in a new populaiton.

## DISCUSSION

In this study we treated the prediction of pneumonia mortality in children as a classification problem. Logistic regression is a widely used methodolgy to model the relationship between a set of predictors and a binary outcome variable, but, in terms of classification, it is often outperformed by other machine learning algorithms such as RF, SVM and ANN. ^9^ These algorithms rely on less assumptions, but at the cost of interpretability and higher risk of overfitting. Given our sparse feature set (16) and our access to a high performance computer, we were able to skip the feature selection process and generate four models (one for each algorithm) for each combination of two or more features. We shortlisted the models that were both parsimonious and predictive, selecting our final model based on the most clincial interpretable features. This final model performed well in both the test and training sets. Overall, our final model has potential to be a useful predictive model for mortality in children under 5 years of age with clinical pneumonia.

As expected, RLR was outperformed by all the other algorithms. While RF and SVM generated more models with AUC ≥ 0.9 than NNET, once we shortlisted the number of models based on the number of features and their measurement reliability we had a similar number of candidate models from each algorithm. This confirms that there is no obvious best algorithm for this type of data, and justifies an approach that initially considers a variety of methods.

There are limited scoring systems or classification methods to identify high-risk children/severity of pneumonia. A formal comparison of our model with these tools was beyond the scope of this paper, but in a recent publication Gallagher et al ^2^ compared the performance of several scores and classification methods and found that the best scoring system (using the number of danger signs from the 2013 World Health Organisation Pocketbook of Hospital Care for Children ^26^) obtained a high AUC (0.82), although the discrete nature of the score did not allow for a threshold that was both sensitive and specific.

## Limitations

Our study has several limitations that could affect the generalisability of our results. First, we noticed that the presence of missing values was more prevalent among children who died than those who survived. If the presence of missing values was related to the severity of the condition the resulting sample after removing cases with missing values would have been healthier than the original one introducing a bias towards less accurate predictions.

Second, we included children in our dataset that met a clinical definition of pneumonia, which was in most cases not confirmed with X-ray, resulting in possible inclusion of children without ‘real’ pneumonia. We believe this can also be viewed as a strength, as it better reflects the reality in low or middle income countries and/or in rural areas.

Third, our study population is representative of a rural area in The Gambia, but there might be notable differences between this and other regions in sub-Saharan Africa due to access to health care and comorbidities such as human immunodeficiency virus, malaria, tuberculosis, and sepsis. Our final model should be used with caution in other populations until its generalizability has been further validated. Alternatively, practitioners should use the methodology described in this paper to create their own local prediction models.

Fourth, during the development of our candidate models we were limited to 16 features that were consistently collected in the Pneumococcal Surveillance Project. Therefore, there could be other better predictors that we may have missed. For example, nasal flaring, grunting, head nodding and vomiting are some features considered in other pneumonia severity scales but that were not among our starting features.

Machine learning algorithms make predictions based on detecting patterns in training data. A weakness of this approach is that it can lead to models with poor interpretability, resulting in counter-intuitive inference. Clinical prediction models are prone to the confounding effects of standard care protocols. We had initially chosen the final model among the shortlisted ones based on what features were included. Despite this model performing well on the test set (AUC= 0.88, sensitivity=0.80 and specificity=0.84), we noted that, leaving all other variables unchanged, an increase in temperature resulted in increasing predicted probability values, which was counter-intuitive for some clinicians we consulted with. It is likely that the algorithm detected this pattern in the data because paracetamol was prescribed to febrile children immediately on admission, which improves their survival rate. Because the acceptability of such counter-intuitive prediction model would be poor, this prompted us to choose a different final model, based on the effect of each variable on the predicted probabilities to avoid counter-intuitive trends and increase the model’s interpretability.

### Strengths

The strengths of our study include its approach to pneumonia mortality in children as a classification problem and its design focused on the generalisability of the final model, not relying only on the performance of the candidate models on the training set, but also taking into account the number of features, their interpretability and reliability. Furthermore, we held out 1/3 of our dataset (with 55 deaths) to test the final model on unseen data.

We addressed the challenges of an imbalanced dataset (very few events) by using three complementary strategies: up and down-sampling (with limited number of up-sampled events), class weighting (penalising the misclassification of events), and using a threshold-invariant performance metric (avoiding the impact of choosing a threshold on a model’s performance during its development).

Despite holding out 1/3 of our dataset and the low incidence of our outcome of interest (only 2 percent deaths), our sample could still be considered large, with more than 10 cases per feature in the training set even for the candidate models with all possible features. Furthermore, the final model, with 5 features, was developed with more than 30 cases per feature.

Another strength of our models was that they predicted the probability of mortality, allowing us to identify the best classification threshold without being limited by discrete values. Finally, since all measurements come from the same study the definitions and procedures are consistent throughout the data collection.

### Implications and conclusion

Our final model showed excellent performance not only in the training set, but also in a previously unseen test set. It could be used to assist with triage decisions. Future research will compare our model with existing models and validate our model on different populations to assess its generalizability.

## Supporting information

Supplemental Files (eTable, eAppendix)

Model .rds file and shiny app

## Data Availability

The data is available on request from Dr. Grant Mackenzie. The code to replicate the results can be found on github: https://github.com/MRCG-djeffries/mortality-prediction.
The model implemented via a Shiny web can be downloaded from the supplemental digital content (as well as github: https://github.com/MRCG-djeffries/mortality-prediction) both as an independent .rds file or as a Shiny App, which incorporates a user interface that allows for individual and batch mortality predictions, as well as validation of our model in new datasets.

## Supplementary Digital Content

Supplemental Digital Content 1. eTable with detailed characteristics of the sample and eAppendix with details of how the final model was selected. (pdf)

Supplemental Digital Content 2. The final model’s rds file and Shiny App (zip)

## Acknowledgements

We thank Drs. Yekini-Ajao Olatunji and Galega Babila Lobga for their assistance in classifying the potential predictors as reliable or less-reliable in terms of their measurement.

## FOOTNOTES

## Abbreviations

AUC: Area Under the Receiver operating characteristic Curve
RF: random forest
SVM: support vector machines
ANN: artificial neural networks

## REFERENCES

1. Liu L, Oza S, Hogan D, et al. Global, regional, and national causes of under-5 mortality in 2000–15: an updated systematic analysis with implications for the Sustainable Development Goals. The Lancet. 2016;388(10063):3027–3035. doi:10.1016/S0140-6736(16)31593-8

2. Gallagher KE, Knoll MD, Prosperi C, et al. The Predictive Performance of a Pneumonia Severity Score in Human Immunodeficiency Virus–negative Children Presenting to Hospital in 7 Low- and Middle-income Countries. Clin Infect Dis. 2020;70(6):1050–1057. doi:10.1093/cid/ciz350

3. Fine MJ, Auble TE, Yealy DM, et al. A Prediction Rule to Identify Low-Risk Patients with Community-Acquired Pneumonia. N Engl J Med. 1997;336(4):243–250. doi:10.1056/NEJM199701233360402

4. Charles PGP, Wolfe R, Whitby M, et al. SMART-COP: A Tool for Predicting the Need for Intensive Respiratory or Vasopressor Support in Community-Acquired Pneumonia. Clin Infect Dis. 2008;47(3):375–384. doi:10.1086/589754

5. Lim WS, van der Eerden MM, Laing R, et al. Defining community acquired pneumonia severity on presentation to hospital: an international derivation and validation study. Thorax. 2003;58(5):377–382. doi:10.1136/thorax.58.5.377

6. Arbo A, Lovera D, Martínez-Cuellar C. Mortality Predictive Scores for Community-Acquired Pneumonia in Children. Curr Infect Dis Rep. 2019;21(3):10. doi:10.1007/s11908-019-0666-9

7. Williams DJ, Zhu Y, Grijalva CG, et al. Predicting Severe Pneumonia Outcomes in Children. PEDIATRICS. 2016;138(4):e20161019–e20161019. doi:10.1542/peds.2016-1019

8. Reed C, Madhi SA, Klugman KP, et al. Development of the Respiratory Index of Severity in Children (RISC) Score among Young Children with Respiratory Infections in South Africa. PLOS ONE. 2012;7(1):e27793. doi:10.1371/journal.pone.0027793

9. Fernandez-Delgado M, Cernadas E, Barro S, Amorim D. Do we Need Hundreds of Classifiers to Solve Real World Classification Problems? J Mach Learn Res. 2014;15:3133–3181.

10. Mackenzie GA, Plumb ID, Sambou S, et al. Monitoring the Introduction of Pneumococcal Conjugate Vaccines into West Africa: Design and Implementation of a Population-Based Surveillance System. PLOS Med. 2012;9(1):e1001161. doi:10.1371/journal.pmed.1001161

11. Kuhn M. Building Predictive Models in R Using the caret Package | Kuhn | Journal of Statistical Software. J Stat Softw. 2008;28(5):1–26. doi:10.18637/jss.v028.i05

12. Breiman L. Random Forests. Mach Learn. 2001;45(1):5–32. doi:10.1023/A:1010933404324

13. Cortes C, Vapnik V. Support-vector networks. Mach Learn. 1995;20(3):273–297. doi:10.1007/BF00994018

14. Ripley BD. Pattern Recognition and Neural Networks. Cambridge University Press; 2007.

15. Lin C-J, Weng RC, Keerthi SS. Trust Region Newton Method for Large-Scale Logistic Regression. J Mach Learn Res. 2008;9:627–650.

16. Moons KGM, Altman DG, Reitsma JB, et al. Transparent Reporting of a multivariable prediction model for Individual Prognosis Or Diagnosis (TRIPOD): Explanation and Elaboration. Ann Intern Med. 2015;162(1):W1. doi:10.7326/M14-0698

17. Peduzzi P, Concato J, Feinstein AR, Holford TR. Importance of events per independent variable in proportional hazards regression analysis II. Accuracy and precision of regression estimates. J Clin Epidemiol. 1995;48(12):1503–1510. doi:10.1016/0895-4356(95)00048-8

18. Peduzzi P, Concato J, Kemper E, Holford TR, Feinstein AR. A simulation study of the number of events per variable in logistic regression analysis. J Clin Epidemiol. 1996;49(12):1373–1379. doi:10.1016/S0895-4356(96)00236-3

19. van der Ploeg T, Austin PC, Steyerberg EW. Modern modelling techniques are data hungry: a simulation study for predicting dichotomous endpoints. BMC Med Res Methodol. 2014;14(1):137. doi:10.1186/1471-2288-14-137

20. Kuhn M. Futility Analysis in the Cross-Validation of Machine Learning Models. ArXiv14056974 Cs Stat. Published online May 27, 2014. Accessed April 16, 2020. http://arxiv.org/abs/1405.6974

21. Kuhn M. 14 Adaptive Resampling | The Caret Package. Accessed January 25, 2021. https://topepo.github.io/caret/adaptive-resampling.html

22. Chawla NV, Bowyer KW, Hall LO, Kegelmeyer WP. SMOTE: Synthetic Minority Over-sampling Technique. J Artif Intell Res. 2002;16:321–357. doi:10.1613/jair.953

23. R Core Team. R: A Language and Environment for Statistical Computing. R Foundation for Statistical Computing; 2020. https://www.R-project.org/

24. Kuhn M. Caret: Classification and Regression Training.; 2020. https://CRAN.R-project.org/package=caret

25. Brownlee J. How to use Learning Curves to Diagnose Machine Learning Model Performance. Machine Learning Mastery. Published February 26, 2019. Accessed September 22, 2020. https://machinelearningmastery.com/learning-curves-for-diagnosing-machine-learning-model-performance/

26. World Health Organization. Pocket Book of Hospital Care for Children: Guidelines for the Management of Common Childhood Illnesses. World Health Organization; 2013.

