## Supplemental Files (eTable, eAppendix) for "DEVELOPMENT AND VALIDATION OF A MODEL FOR THE PREDICTION OF MORTALITY IN CHILDREN UNDER FIVE YEARS WITH CLINICAL PNEUMONIA IN RURAL GAMBIA"

**eTable 1. Detailed Characteristics of the Sample.**

| Variable | Descriptives before inclusion criteria | Descriptives after inclusion criteria | Descriptives before removing *missings* All | Descriptives before removing *missings* Non-survivors | Descriptives before removing *missings* Survivors | Descriptives after removing *missings* |
|---|---|---|---|---|---|---|
| **Mortality** | | | | | | |
| No | 15048 (97.4%) | 11419 (97.6%) | 11419 (97.6%) | 0 (0%) | 11419 (100%) | 10790 (98%) |
| Yes | 408 (2.6%) | 279 (2.4%) | 279 (2.4%) | 279 (100%) | 0 (0%) | 222 (2%) |
| *missing* | 0 (0%) | 0 (0%) | 0 (0%) | 0 (0%) | 0 (0%) | 0 (0%) |
| **Age** (days)[a] | 363 days (181, 674) | 348 days (166.2, 636) | 348 days (166.2, 636) | 317 days (132, 674) | 349 days (168.5, 635) | 348 days (169, 633) |
| *missing* | 0 (0%) | 0 (0%) | 0 (0%) | 0 (0%) | 0 (0%) | 0 (0%) |
| **Sex** (% Male)[a] | | | | | | |
| Female | 6934 (44.9%) | 5296 (45.3%) | 5296 (45.3%) | 144 (51.6%) | 5152 (45.1%) | 4997 (45.4%) |
| Male | 8521 (55.1%) | 6401 (54.7%) | 6401 (54.7%) | 135 (48.4%) | 6266 (54.9%) | 6015 (54.6%) |
| *missing* | 1 (0%) | 1 (0%) | 1 (0%) | 0 (0%) | 1 (0%) | 0 (0%) |
| **Inability to drink or breastfeed** (% yes)[b] | | | | | | |
| No | 13681 (88.5%) | 10432 (89.2%) | 10432 (89.2%) | 154 (55.2%) | 10278 (90%) | 9845 (89.4%) |
| Yes | 1466 (9.5%) | 1241 (10.6%) | 1241 (10.6%) | 125 (44.8%) | 1116 (9.8%) | 1167 (10.6%) |
| *missing* | 309 (2%) | 25 (0.2%) | 25 (0.2%) | 0 (0%) | 25 (0.2%) | 0 (0%) |
| **Inability to sit unsupported** (% yes)[a] | | | | | | |
| No | 14585 (94.4%) | 11290 (96.5%) | 11290 (96.5%) | 194 (69.5%) | 11096 (97.2%) | 10720 (97.3%) |
| Yes | 509 (3.3%) | 342 (2.9%) | 342 (2.9%) | 80 (28.7%) | 262 (2.3%) | 292 (2.7%) |
| *missing* | 362 (2.3%) | 66 (0.6%) | 66 (0.6%) | 5 (1.8%) | 61 (0.5%) | 0 (0%) |
| **Convulsions** (% yes)[a] | | | | | | |
| No | 14493 (93.8%) | 11273 (96.4%) | 11273 (96.4%) | 231 (82.8%) | 11042 (96.7%) | 10636 (96.6%) |
| Yes | 691 (4.5%) | 414 (3.5%) | 414 (3.5%) | 47 (16.8%) | 367 (3.2%) | 376 (3.4%) |
| *missing* | 272 (1.8%) | 11 (0.1%) | 11 (0.1%) | 1 (0.4%) | 10 (0.1%) | 0 (0%) |
| **Lethargy** (% yes)[b] | | | | | | |

| Variable | Descriptives before inclusion criteria | Descriptives after inclusion criteria | Descriptives before removing *missing*s All | Descriptives before removing *missing*s Non-survivors | Descriptives before removing *missing*s Survivors | Descriptives after removing *missing*s |
|---|---|---|---|---|---|---|
| No | 12647 (81.8%) | 9885 (84.5%) | 9885 (84.5%) | 88 (31.5%) | 9797 (85.8%) | 9384 (85.2%) |
| Yes | 2512 (16.3%) | 1790 (15.3%) | 1790 (15.3%) | 189 (67.7%) | 1601 (14%) | 1628 (14.8%) |
| *missing* | 297 (1.9%) | 23 (0.2%) | 23 (0.2%) | 2 (0.7%) | 21 (0.2%) | 0 (0%) |
| **Lower chest wall indrawing** (% yes)[b] | | | | | | |
| No | 9570 (61.9%) | 6088 (52%) | 6088 (52%) | 147 (52.7%) | 5941 (52%) | 5817 (52.8%) |
| Yes | 5624 (36.4%) | 5605 (47.9%) | 5605 (47.9%) | 130 (46.6%) | 5475 (47.9%) | 5195 (47.2%) |
| *missing* | 262 (1.7%) | 5 (0%) | 5 (0%) | 2 (0.7%) | 3 (0%) | 0 (0%) |
| **Wheeze** (% yes)[b] | | | | | | |
| No | 11756 (76.1%) | 8529 (72.9%) | 8529 (72.9%) | 238 (85.3%) | 8291 (72.6%) | 8097 (73.5%) |
| Yes | 3432 (22.2%) | 3160 (27%) | 3160 (27%) | 39 (14%) | 3121 (27.3%) | 2915 (26.5%) |
| *missing* | 268 (1.7%) | 9 (0.1%) | 9 (0.1%) | 2 (0.7%) | 7 (0.1%) | 0 (0%) |
| **Vaccination status** (% vaccinated)[b, c] | | | | | | |
| Not vaccinated | 5433 (35.2%) | 4217 (36%) | 4217 (36%) | 122 (43.7%) | 4095 (35.9%) | 3687 (33.5%) |
| Vaccinated | 10023 (64.8%) | 7481 (64%) | 7481 (64%) | 157 (56.3%) | 7324 (64.1%) | 7325 (66.5%) |
| *missing* | 0 (0%) | 0 (0%) | 0 (0%) | 0 (0%) | 0 (0%) | 0 (0%) |
| **Axillary temperature** (°C)[a] | 37.9 (37, 38.8) | 38 (37.1, 38.8) | 38 (37.1, 38.8) | 37.7 (36.7, 38.5) | 38 (37.1, 38.8) | 37.9 (37.1, 38.8) |
| *missing* | 39 (0.3%) | 9 (0.1%) | 9 (0.1%) | 1 (0.4%) | 8 (0.1%) | 0 (0%) |
| **Number of days unwell** [b, d] | 3 (2, 4) | 3 (2, 4) | 3 (2, 4) | 3 (3, 6) | 3 (2, 3) | 3 (2, 4) |
| *missing* | 406 (2.6%) | 55 (0.5%) | 55 (0.5%) | 9 (3.2%) | 46 (0.4%) | 0 (0%) |
| **Respiratory rate** (breaths/minute)[a] | 52 breaths/min (44, 60) | 56 breaths/min (49, 62) | 56 breaths/min (49, 62) | 54 breaths/min (42, 66) | 56 breaths/min (49, 62) | 56 breaths/min (49, 62) |
| *missing* | 266 (1.7%) | 23 (0.2%) | 23 (0.2%) | 2 (0.7%) | 21 (0.2%) | 0 (0%) |

| Variable | Descriptives before inclusion criteria | Descriptives after inclusion criteria | Descriptives before removing *missing*s<br>All | Descriptives before removing *missing*s<br>Non-survivors | Descriptives before removing *missing*s<br>Survivors | Descriptives after removing *missing*s |
|---|---|---|---|---|---|---|
| **Heart rate** (beats/minute)[a] | 152 beats/min (138, 166) | 155 beats/min (141, 168) | 155 beats/min (141, 168) | 162 beats/min (133, 180) | 155 beats/min (142, 168) | 155 beats/min (142, 168) |
| *missing* | 253 (1.6%) | 0 (0%) | 0 (0%) | 0 (0%) | 0 (0%) | 0 (0%) |
| **Weight-for-height z-score**[a, e] | -1.2 (-2.2, -0.3) | -1.2 (-2.1, -0.2) | -1.2 (-2.1, -0.2) | -2.5 (-3.7, -1.1) | -1.2 (-2.1, -0.2) | -1.2 (-2.1, -0.2) |
| *missing* | 273 (1.8%) | 159 (1.4%) | 159 (1.4%) | 31 (11.1%) | 128 (1.1%) | 0 (0%) |
| **Mid-Upper Arm Circumference** (cm)[a] | 13.5 cm (12.5, 14.4) | 13.5 cm (12.5, 14.4) | 13.5 cm (12.5, 14.4) | 11.5 cm (10.2, 13.2) | 13.5 cm (12.5, 14.4) | 13.5 cm (12.5, 14.4) |
| *missing* | 64 (0.4%) | 33 (0.3%) | 33 (0.3%) | 4 (1.4%) | 29 (0.3%) | 0 (0%) |
| **Oxygen saturation** (%) | 98% (96, 100) | 98% (95, 99) | 98% (95, 99) | 97% (91, 100) | 98% (95, 99) | 98% (95, 99) |
| *missing* | 641 (4.1%) | 358 (3.1%) | 358 (3.1%) | 6 (2.2%) | 352 (3.1%) | 0 (0%) |
| Weight (kg) | 7.8 kg (6.4, 9.6) | 7.8 kg (6.3, 9.4) | | | | |
| *missing* | 43 (0.3%) | 12 (0.1%) | | | | |
| Height (cm) | 72.3 cm (65, 81) | 71.8 cm (64.5, 80.2) | | | | |
| *missing* | 112 (0.7%) | 60 (0.5%) | | | | |
| Malnutrition: Weight-for-age z-core[e] | -1.3 (-2.2, -0.3) | -1.2 (-2.2, -0.3) | | | | |
| *missing* | 166 (1.1%) | 95 (0.8%) | | | | |
| Haemoglobin (g/dL) | 7.3 (1.1, 10.8) | 5.1 (1.1, 10.7) | 5.1 (1.1, 10.7) | 8.2 (1.5, 11) | 5 (1.1, 10.7) | |
| *missing* | 13411 (86.8%) | 10367 (88.6%) | 10367 (88.6%) | 210 (75.3%) | 10157 (88.9%) | |
| Cough (% yes) | | | | | | |
| No | 872 (5.6%) | 127 (1.1%) | | | | |
| Yes | 5363 (34.7%) | 4714 (40.3%) | | | | |
| *missing* | 9221 (59.7%) | 6857 (58.6%) | | | | |
| Difficulty breathing / Shortness of breath (% yes) | | | | | | |

| Variable | Descriptives before inclusion criteria | Descriptives after inclusion criteria | Descriptives before removing *missings* All | Descriptives before removing *missings* Non-survivors | Descriptives before removing *missings* Survivors | Descriptives after removing *missings* |
|---|---|---|---|---|---|---|
| No | 4899 (31.7%) | 2612 (22.3%) | | | | |
| Yes | 10335 (66.9%) | 9083 (77.6%) | | | | |
| *missing* | 222 (1.4%) | 3 (0%) | | | | |
| Nasal Flaring (% yes) | | | | | | |
| No | 9042 (58.5%) | 6316 (54%) | | | | |
| Yes | 5327 (34.5%) | 4792 (41%) | | | | |
| *missing* | 1087 (7%) | 590 (5%) | | | | |
| Grunting (% yes) | | | | | | |
| No | 12171 (78.7%) | 9134 (78.1%) | | | | |
| Yes | 2198 (14.2%) | 1974 (16.9%) | | | | |
| *missing* | 1087 (7%) | 590 (5%) | | | | |
| Abnormal conscious state | | | | | | |
| Alert | 14602 (94.5%) | 11303 (96.6%) | | | | |
| Responds to pain | 195 (1.3%) | 132 (1.1%) | | | | |
| Responds to Voice | 171 (1.1%) | 112 (1%) | | | | |
| Unresponsive | 70 (0.5%) | 46 (0.4%) | | | | |
| *missing* | 418 (2.7%) | 105 (0.9%) | | | | |
| Diarrhoea (% yes) | | | | | | |
| No | 11409 (73.8%) | 8896 (76%) | | | | |
| Yes | 3601 (23.3%) | 2447 (20.9%) | | | | |
| *missing* | 446 (2.9%) | 355 (3%) | | | | |
| Malaria (% yes) | | | | | | |
| No | 10758 (69.6%) | 8550 (73.1%) | | | | |
| Yes | 673 (4.4%) | 465 (4%) | | | | |
| *missing* | 4025 (26%) | 2683 (22.9%) | | | | |

| Variable | Descriptives before inclusion criteria | Descriptives after inclusion criteria | Descriptives before removing *missings*<br>All | Descriptives before removing *missings*<br>Non-survivors | Descriptives before removing *missings*<br>Survivors | Descriptives after removing *missings* |
|---|---|---|---|---|---|---|
| Meningitis (% yes) | | | | | | |
| Yes | 16 (0.1%) | 13 (0.1%) | | | | |
| *missing* | 15440 (99.9%) | 11685 (99.9%) | | | | |
| Bacteraemia (% yes) | | | | | | |
| No | 10 (0.1%) | 9 (0.1%) | | | | |
| Yes | 437 (2.8%) | 341 (2.9%) | | | | |
| *missing* | 15009 (97.1%) | 11348 (97%) | | | | |
| Clinical pneumonia (% yes) | | | | | | |
| No | 3758 (24.3%) | 0 (0%) | | | | |
| Yes | 11698 (75.7%) | 11698 (100%) | | | | |
| *missing* | 0 (0%) | 0 (0%) | | | | |
| Radiological pneumonia (% yes) | | 10408 (89%) | | | | |
| No | 14060 (91%) | 1290 (11%) | | | | |
| Yes | 1396 (9%) | 0 (0%) | | | | |
| *missing* | 0 (0%) | | | | | |

All categorical variables were binary. Continuous outcomes are reported as median (inter-quartile range). Abnormal conscious state and Hamoglobin were excluded.

[a] Considered by a panel of three clinicians to be a reliably measured variable.

[b] Considered by a panel of three clinicians to be a variable measured in a less reliable way (subjective and/or skill-dependent)

[c] Considered not vaccinated if the patient had not received neither the second nor the third dose of the pneumococcal vaccine.

[d] Number of days the patient had been feeling unwell before admission.

[e] Calculated using the World Health Organisation reference tables ('anthro' package)

**eAppendix: Selection of the final model details.**

We had initially chosen the final model based on the included features, with a clinical perspective in mind. This initial final model had been generated by NNET (neural network) using the variables age, axillary temperature, heart rate, mid-upper arm circumference, oxygen saturation and history of convulsions. When we applied our final model to the test set for its validation, the AUC was 0.87 (95% confidence interval: 0.82 to 0.92) and, when we classified subjects in the test set applying the threshold probability 0.67 (the best threshold value identified during the development of the model using the trianign set), the resulting sensitivity and specificity were 0.80 and 0.84, respectively.

Despite this model's excellent performance when we validated it on the test set, the effect of some variables on the predicted probabilities was counter-intuitive. Specifically, increasing temperature values resulted in decreasing predicted probabilities of death when all other variables remained the same (eFigure 1).

**eFigure 1: Changes in the predicted probability of the initially selected final model when modifying each variable while leaving the rest at a reference value**

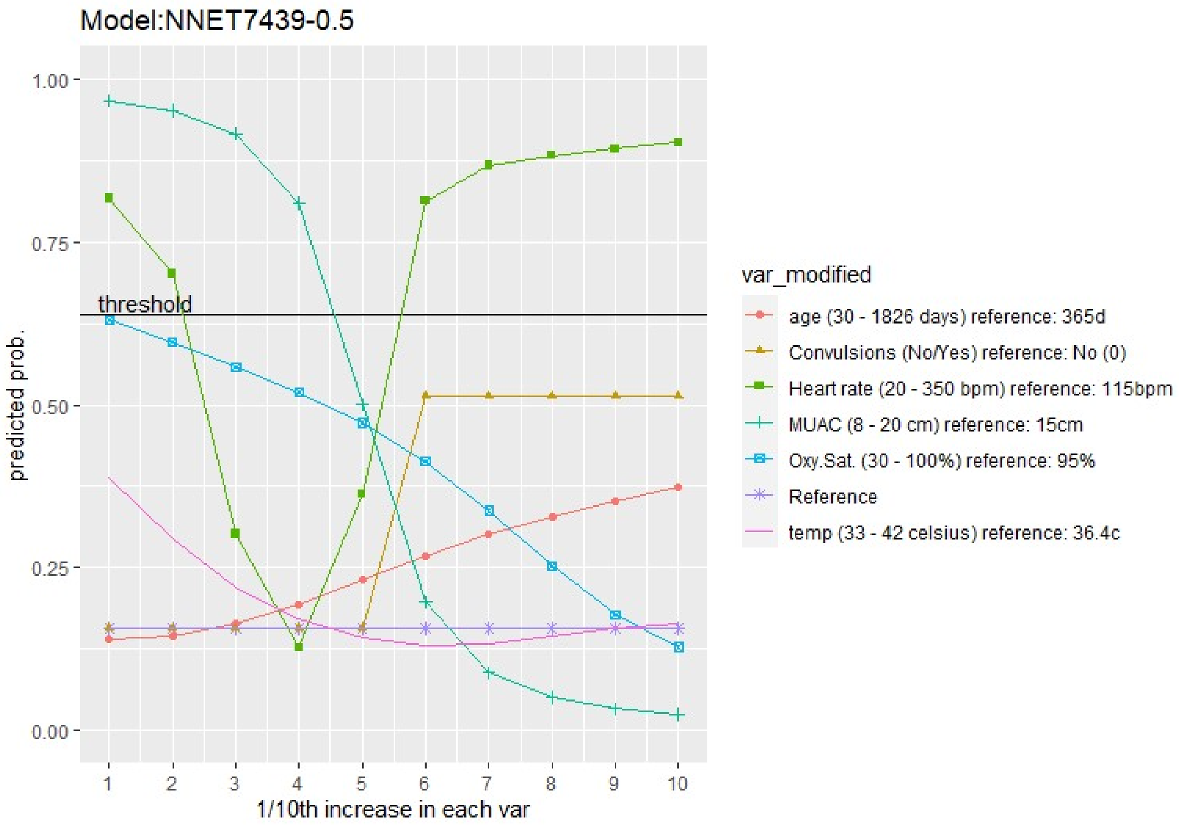


This was counter-intuitive for some clinicians we presented the model to and, because the acceptability of such a counter-intuitive prediction model would be poor, we decided to re-assess all of the nine shortlisted models and decide on the final model based on the effect of each variable on the predicted probabilities to avoid counter-intuitive trends and increase the model's interpretability.

eFigure 2 provides a summary of the shortlisted models and Figures S3 to S10 show the changes in the predicted probability of each model when modifying each variable while leaving the rest at a reference value. The threshold line indicates the best classification threshold for that model in the training set. It is important to note that all these values were obtained on the training set. The test set was only used after a decision was made on the new final model (RF3436-1.00).

**eFigure 2: Summary of the characteristics and performance (on the training set) of all shortlisted models. Model 2 was the one initially selected. Model 8 is the current final model.**

| # | model | feat.comb | n.feats | Weight | AUC | best thresh | Sens thresh | Spec thresh | features |
|---|---|---|---|---|---|---|---|---|---|
| 1 | SVM | 4454.00 | 5.00 | 0.67 | 0.86 | 0.49 | 0.80 | 0.80 | Temp, Heart.Rate, MUAC, Inability.Sit, Convulsions |
| 2 | NNET | 7439.00 | 6.00 | 0.50 | 0.86 | 0.64 | 0.81 | 0.80 | age, Temp, Heart.Rate, MUAC, Oxy.Sat, Convulsions |
| 3 | RF | 11231.00 | 6.00 | 1.00 | 0.87 | 0.43 | 0.81 | 0.81 | Temp, Heart.Rate, MUAC, Oxy.Sat, Inability.Sit, Convulsions |
| 4 | SVM | 3458.00 | 5.00 | 0.50 | 0.90 | 0.59 | 0.83 | 0.84 | age, Heart.Rate, MUAC, Convulsions, Lethargy* |
| 5 | SVM | 4459.00 | 5.00 | 0.25 | 0.90 | 0.47 | 0.87 | 0.80 | Temp, Heart.Rate, MUAC, Convulsions, Lethargy* |
| 6 | NNET | 2676.00 | 5.00 | 1.00 | 0.90 | 0.45 | 0.84 | 0.82 | age, Temp, Heart.Rate, MUAC, Lethargy* |
| 7 | NNET | 3458.00 | 5.00 | 1.00 | 0.90 | 0.43 | 0.83 | 0.80 | age, Heart.Rate, MUAC, Convulsions, Lethargy* |
| 8 | RF | 3436.00 | 5.00 | 1.00 | 0.90 | 0.44 | 0.83 | 0.83 | age, Heart.Rate, MUAC, Oxy.Sat, Lethargy* |
| 9 | RF | 3454.00 | 5.00 | 1.00 | 0.90 | 0.47 | 0.82 | 0.84 | age, Heart.Rate, MUAC, Inability.Sit, Lethargy* |

**eFigure 3: Changes in the predicted probability of shortlisted model 1 when modifying each variable while leaving the rest at a reference value.**

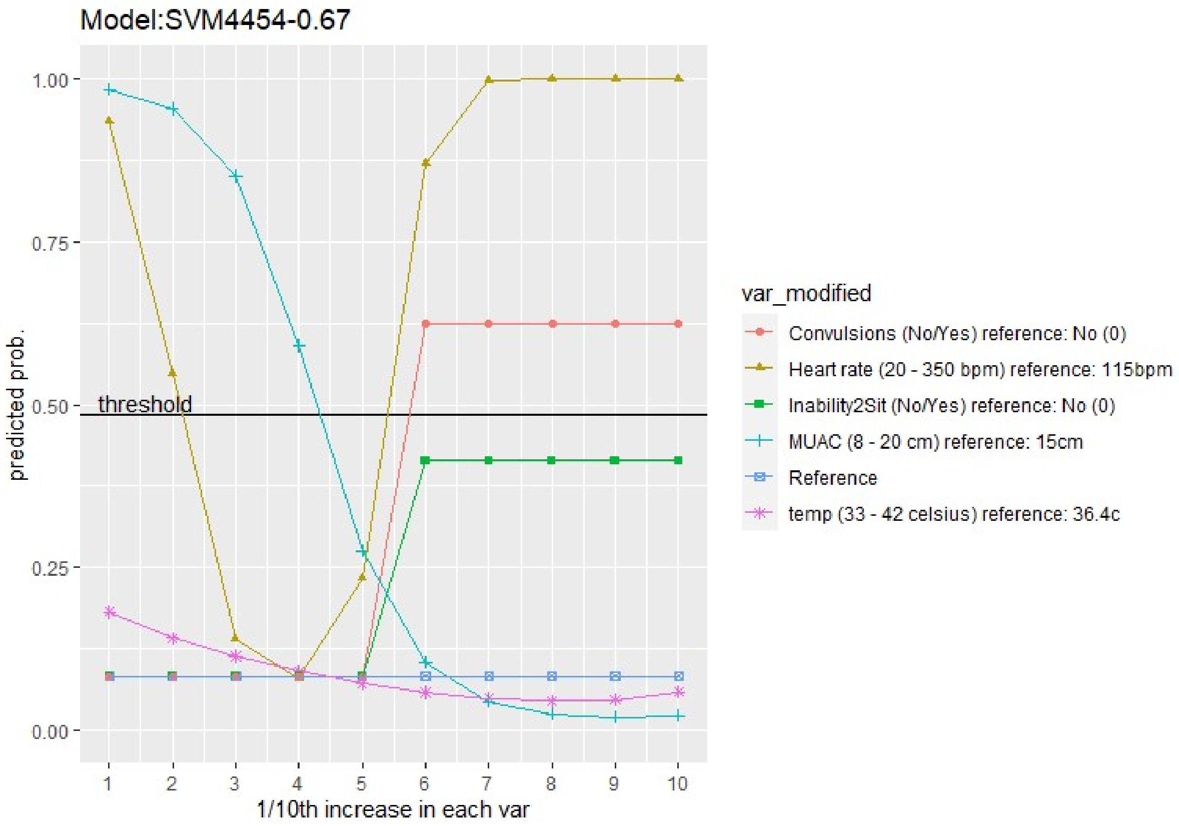

**eFigure 4: Changes in the predicted probability of shortlisted model 3 when modifying each variable while leaving the rest at a reference value.**

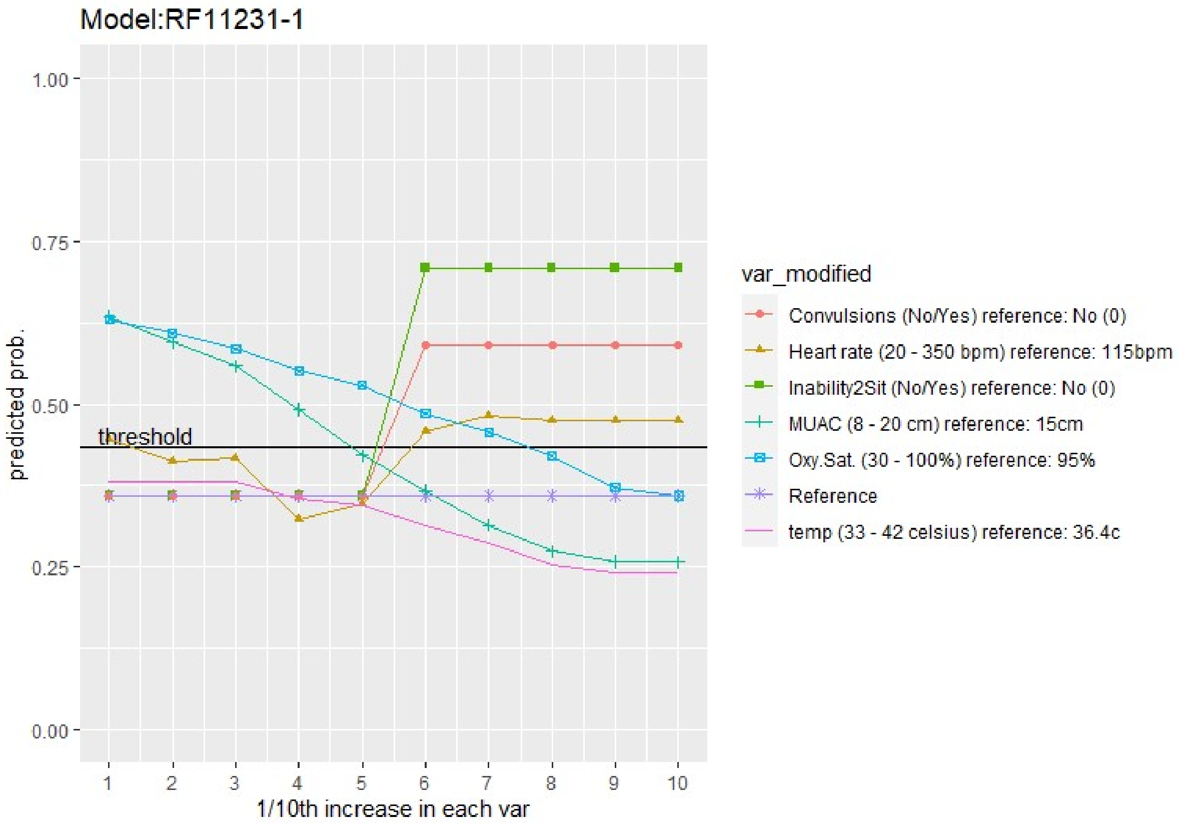


**eFigure 5: Changes in the predicted probability of shortlisted model 4 when modifying each variable while leaving the rest at a reference value.**

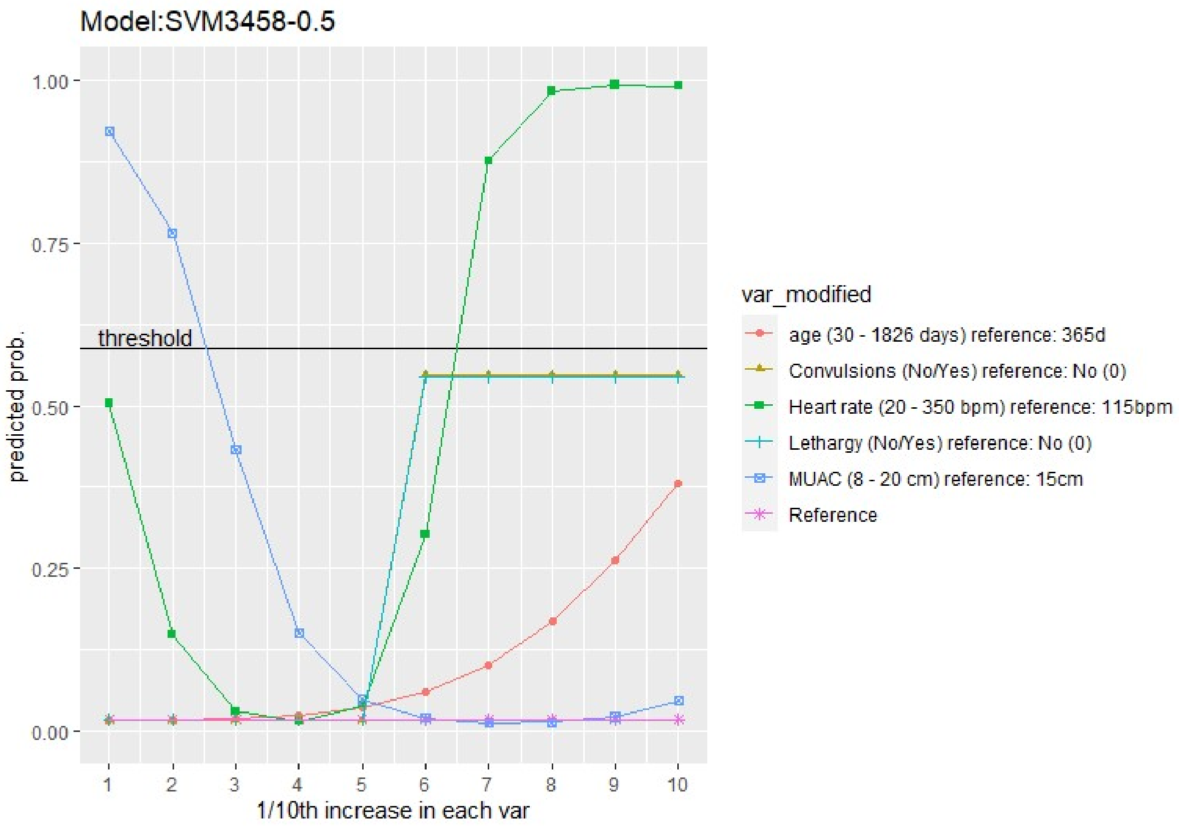


**eFigure 6: Changes in the predicted probability of shortlisted model 5 when modifying each variable while leaving the rest at a reference value.**

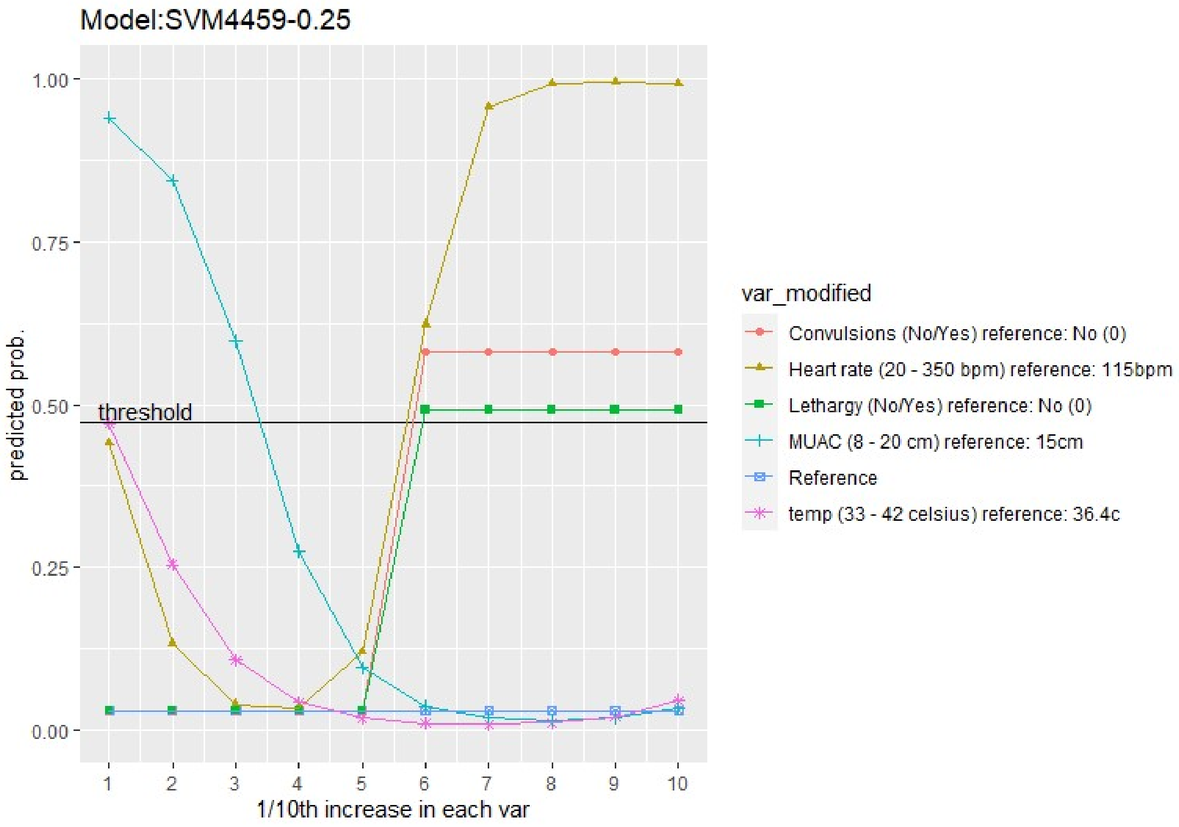

**eFigure 7: Changes in the predicted probability of shortlisted model 6 when modifying each variable while leaving the rest at a reference value.**

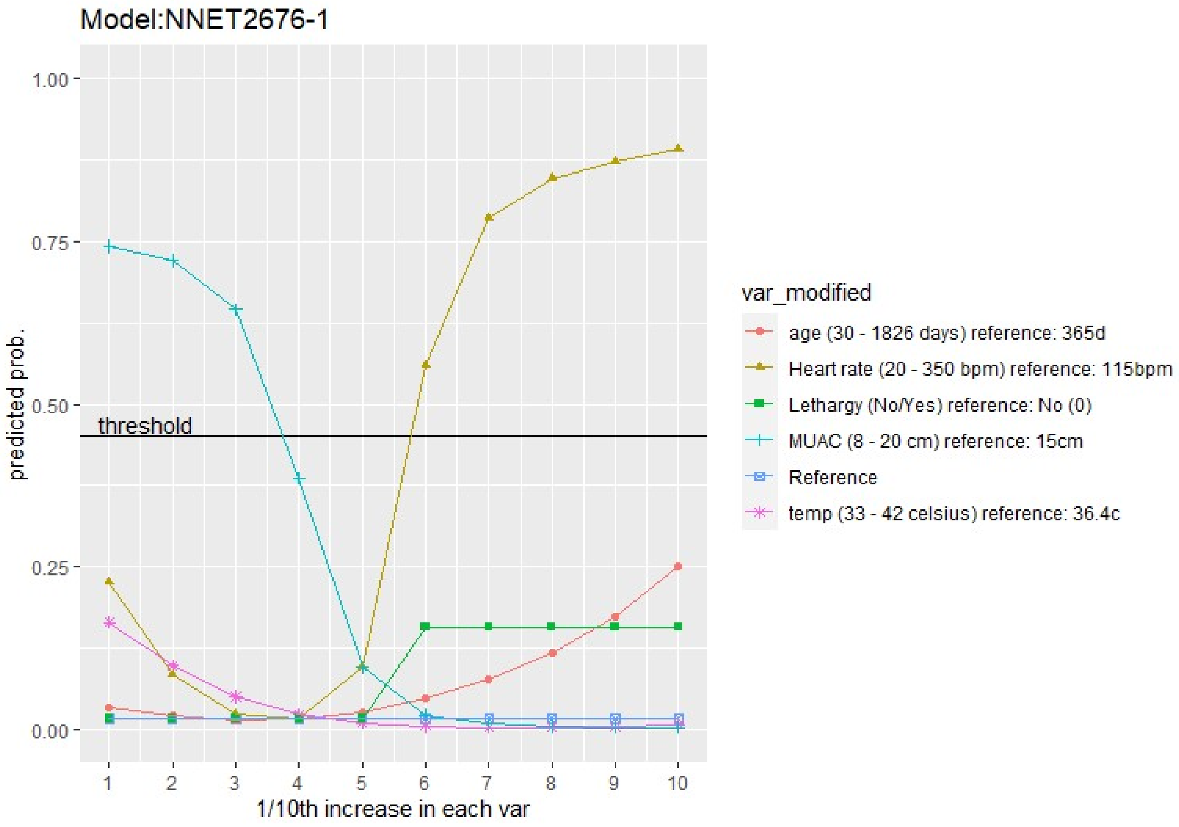


**eFigure 8: Changes in the predicted probability of shortlisted model 7 when modifying each variable while leaving the rest at a reference value.**

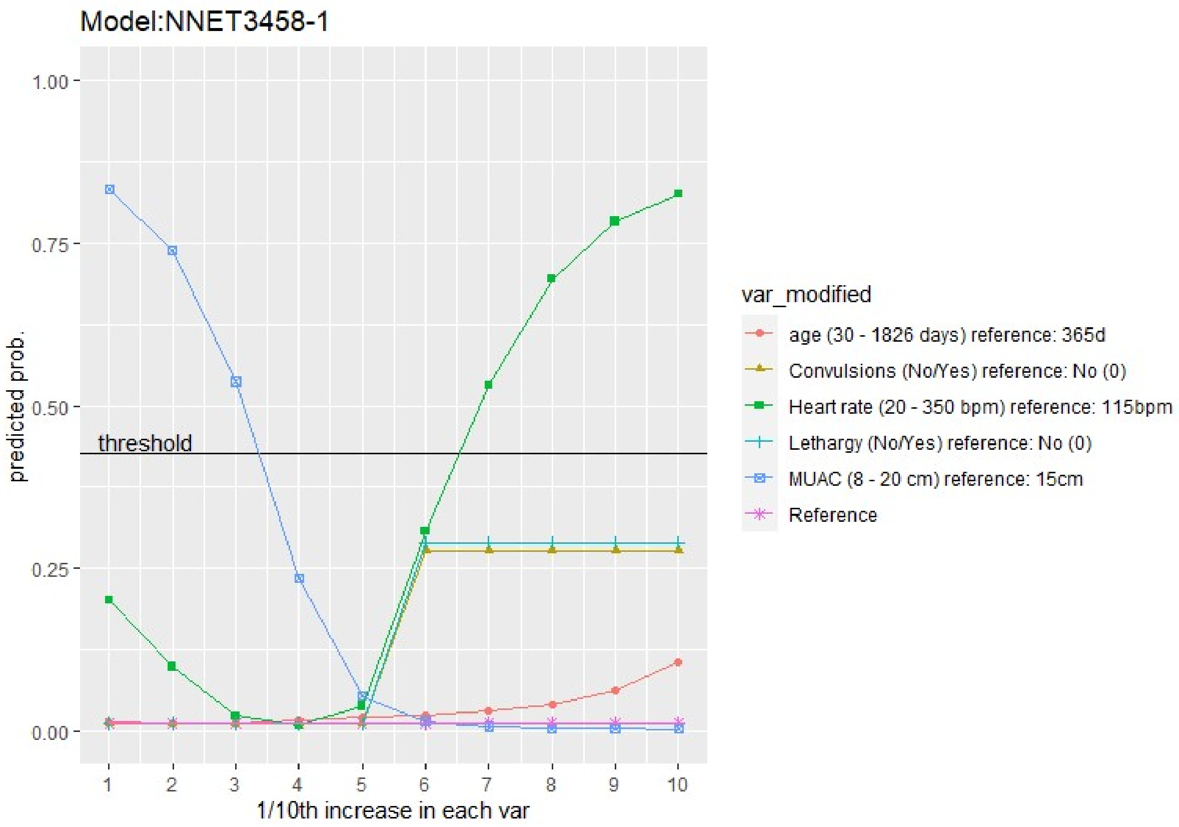


**eFigure 9: Changes in the predicted probability of shortlisted model 8 when modifying each variable while leaving the rest at a reference value.**

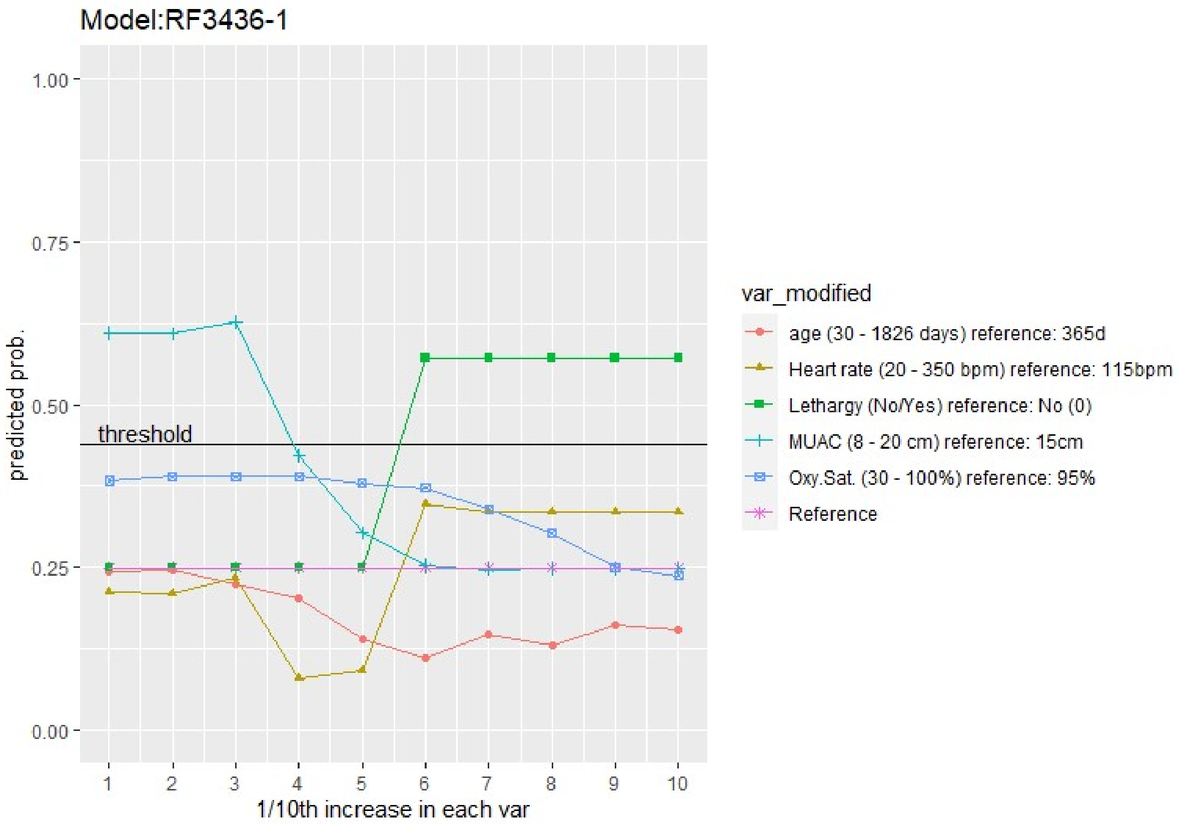

**eFigure 10: Changes in the predicted probability of shortlisted model 9 when modifying each variable while leaving the rest at a reference value.**

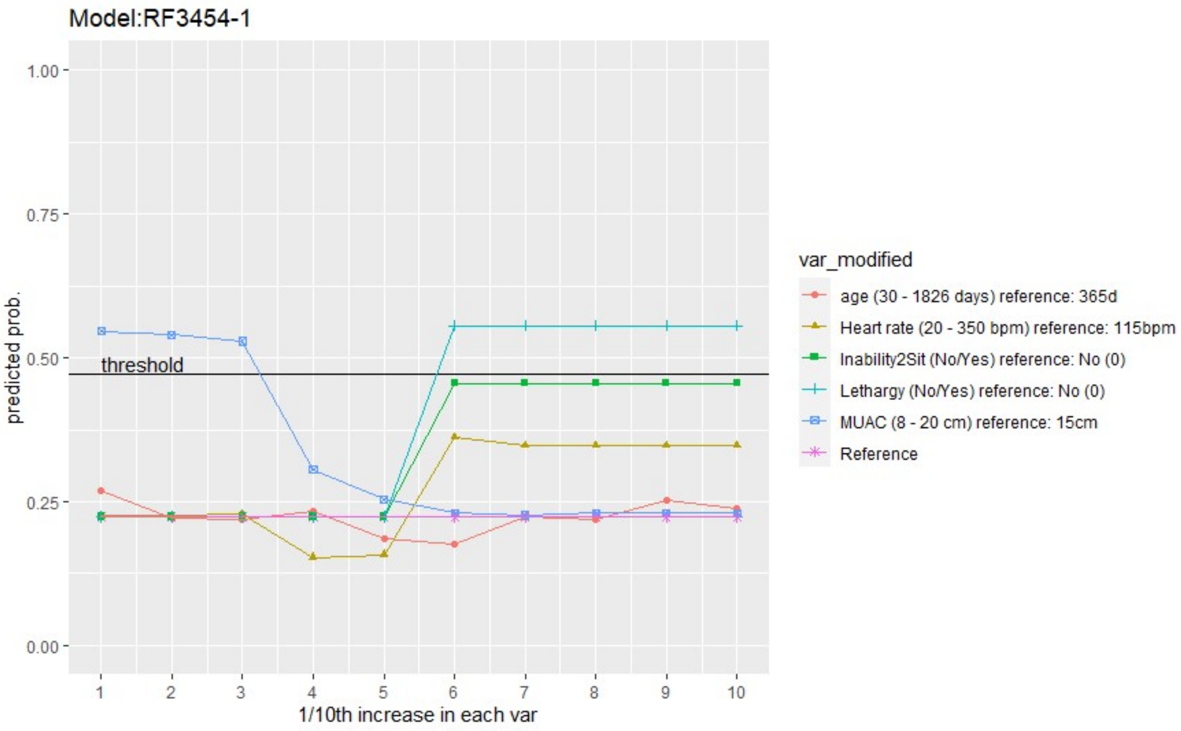
