## Supplementary material for "DEVELOPMENT AND VALIDATION OF A MODEL FOR THE PREDICTION OF MORTALITY IN CHILDREN UNDER FIVE YEARS WITH CLINICAL PNEUMONIA IN RURAL GAMBIA": Model .rds file and shiny app: app.R

```
library(shiny)
library(caret)
library(data.table)
library(ggplot2)

#options(shiny.maxRequestSize = 30*1024^2)


draw_confusion_matrix <- function(cm) {
  ### Copied and modified from https://stackoverflow.com/a/53235386
  
  total <- sum(cm$table)
  res <- as.numeric(cm$table)
  
  # Generate color gradients. Palettes come from RColorBrewer.
  greenPalette <- c("#F7FCF5","#E5F5E0","#C7E9C0","#A1D99B","#74C476","#41AB5D","#238B45","#006D2C","#00441B")
  redPalette <- c("#FFF5F0","#FEE0D2","#FCBBA1","#FC9272","#FB6A4A","#EF3B2C","#CB181D","#A50F15","#67000D")
  getfontColor <- function (greenOrRed = "green", amount = 0) {
    if (amount == 0)
      return("#000000")
    palette <- c("gray10","gray20","gray30","gray40","gray50","gray60","gray70","gray80","gray90")
    colorRampPalette(palette)(100)[10 + ceiling(90 * amount / total)]
  }
  getColor <- function (greenOrRed = "green", amount = 0) {
    if (amount == 0)
      return("#FFFFFF")
    palette <- greenPalette
    if (greenOrRed == "red")
      palette <- redPalette
    colorRampPalette(palette)(100)[10 + ceiling(90 * amount / total)]
  }
  
  # set the basic layout
  layout(matrix(c(1,1,2)))
  par(mar=c(2,2,2,2))
  plot(c(100, 345), c(300, 450), type = "n", xlab="", ylab="", xaxt='n', yaxt='n')
  title('CONFUSION MATRIX', cex.main=2)
  
  # create the matrix 
  classes = colnames(cm$table)
  rect(150, 430, 240, 370, col=getColor("green", res[1]))
  text(195, 435, classes[1], cex=1.2)
  rect(250, 430, 340, 370, col=getColor("red", res[3]))
  text(295, 435, classes[2], cex=1.2)
  text(125, 370, 'Predicted', cex=1.3, srt=90, font=2)
  text(245, 450, 'Actual', cex=1.3, font=2)
  rect(150, 305, 240, 365, col=getColor("red", res[2]))
  rect(250, 305, 340, 365, col=getColor("green", res[4]))
  text(140, 400, classes[1], cex=1.2, srt=90)
  text(140, 335, classes[2], cex=1.2, srt=90)
  
  # prepare the cm results
  det11a <- paste0(round(res[1]/(res[1]+res[2])*100,1), "% of all actually ", classes[1], "   ")
  det11b <- paste0(round(res[1]/(res[1]+res[3])*100,1), "% of all predicted ", classes[1])
  det22a <- paste0(round(res[4]/(res[4]+res[3])*100,1), "% of all actually ", classes[2], "   ")
  det22b <- paste0(round(res[4]/(res[4]+res[2])*100,1), "% of all predicted ", classes[2])
  
  # add in the cm results
  # text(195, 400, res[1], cex=1.6, font=2, col='gray5')
  # text(295, 335, res[4], cex=1.6, font=2, col='gray20')
  fontcolor <- "gray25"
  text(195, 407, res[1], cex=1.6, font=2, col=getfontColor("green", res[1]))
    text(195, 397, det11a, cex=1.2, font=1, col=getfontColor("green", res[1]))
    text(195, 389, det11b, cex=1.2, font=1, col=getfontColor("green", res[1]))
  text(195, 335, res[2], cex=1.6, font=2, col=getfontColor("red", res[2]))
  text(295, 400, res[3], cex=1.6, font=2, col=getfontColor("red", res[3]))
  text(295, 342, res[4], cex=1.6, font=2, col=getfontColor("green", res[4]))
    text(295, 332, det22a, cex=1.2, font=1, col=getfontColor("green", res[4]))
    text(295, 324, det22b, cex=1.2, font=1, col=getfontColor("green", res[4]))
  
  # add in the specifics 
  plot(c(100, 0), c(100, 0), type = "n", xlab="", ylab="", main = "DETAILS", xaxt='n', yaxt='n')
  text(10, 85, names(cm$overall[1]), cex=1.2, font=2)
  text(10, 70, round(as.numeric(cm$overall[1]), 3), cex=1.2)
  text(30, 85, names(cm$overall[2]), cex=1.2, font=2)
  text(30, 70, round(as.numeric(cm$overall[2]), 3), cex=1.2)
  text(50, 85, names(cm$byClass[5]), cex=1.2, font=2)
  text(50, 70, round(as.numeric(cm$byClass[5]), 3), cex=1.2)
  text(70, 85, names(cm$byClass[6]), cex=1.2, font=2)
  text(70, 70, round(as.numeric(cm$byClass[6]), 3), cex=1.2)
  text(90, 85, names(cm$byClass[7]), cex=1.2, font=2)
  text(90, 70, round(as.numeric(cm$byClass[7]), 3), cex=1.2)
  
  # add in the accuracy information 
  text(30, 35, names(cm$byClass[1]), cex=1.5, font=2)
  text(30, 20, round(as.numeric(cm$byClass[1]), 3), cex=1.4)
  text(70, 35, names(cm$byClass[2]), cex=1.5, font=2)
  text(70, 20, round(as.numeric(cm$byClass[2]), 3), cex=1.4)
}


# Define UI for data upload app ----
ui <- fluidPage(
  
  # App title ----
  titlePanel("MRC 2020"),
  
  # Sidebar layout with input and output definitions ----
  sidebarLayout(
    
    
    # Sidebar panel for inputs ----
    sidebarPanel(
      # Input: One or many
      radioButtons(inputId="choice", label = "What do you want to do?",
                   choices = list("Prediction for one case" = "one", 
                                  "Predictions for many (dataset)" = "many",
                                  "Validation in new dataset" = "validation"), 
                   selected = "many"),
      
      # Starts file loader (if choice is 'many' or 'validation')
      conditionalPanel(condition = "(input.choice == 'validation' | input.choice == 'many')",
                         tags$hr(),
                         
                         # Input: Select a file ----
                         fileInput(inputId = "dataset", 
                                   label = "Select CSV file with your data",
                                   multiple = FALSE,
                                   accept = c("text/csv",
                                              "text/comma-separated-values,text/plain",
                                              ".csv")),
                         # Horizontal line ----
                         tags$hr(),
                         
                         # If dataset is loaded
                         conditionalPanel(
                           condition = "output.fileUploaded",
                           #strong("Please specify the column names identifying the following variables:"),
                           # Input: Name of appropriate headers
                           selectInput(inputId = "input_Age_name", label="a", choices = ""),
                           selectInput(inputId = "input_HR_name", label="a", choices = ""),
                           selectInput(inputId = "input_MUAC_name", label="a", choices = ""),
                           selectInput(inputId = "input_OxySat_name", label="a", choices = ""),
                           selectInput(inputId = "input_Lethargy_name", label="a", choices = ""),
                           conditionalPanel(condition = "input.input_Lethargy_name!=''",
                                            p("Select the value that codes for..."),
                                            selectInput(inputId = "lethNoLabel", label="No", choices = "", width = '70%'),
                                            selectInput(inputId = "lethYesLabel", label="Yes", choices = "", width = '70%')
                           ),
                           # If choice is 'validation'
                           conditionalPanel(condition = "input.choice == 'validation'",
                                              selectInput(inputId = "input_Death_name", label = "a", choices = ""),
                                              conditionalPanel(condition = "input.input_Death_name!=''",
                                                              p("Select the value that codes for..."),
                                                              selectInput(inputId = "deathNoLabel", label="No", choices = "", width = '70%'),
                                                              selectInput(inputId = "deathYesLabel", label="Yes", choices = "", width = '70%')
                                           
                                              )
                           )
                         ) # Finishes condition if dataset is loaded
                       
                       ), # Finishes file loader (if choice is 'many' or 'validation')
      
      

######################

      # If choice is 'one'
      conditionalPanel(condition = "input.choice == 'one'",
                       tags$hr(),
                       
                         # Input: Name of appropriate headers
                         sliderInput(inputId = "input_Age", label="Age (in days)", min=30, max=1826, value=365, step=1),
                         sliderInput(inputId = "input_HR", label="Heart rate (in beats/min)", min=20, max=350, value=100, step=1),
                         sliderInput(inputId = "input_MUAC", label="MUAC (in cm)", min=8, max=20, value=20, step=0.1),
                         sliderInput(inputId = "input_OxySat", label="Oxygen saturation (in %)", min=30, max=100, value=100, step=1),
                         radioButtons(inputId = "input_Lethargy", label="Lethargy", choices = list("No"=0, "Yes"=1), inline=TRUE),
                         actionButton(inputId = "input_submit", label = "Calculate probability value")
      ), # Finishes if choice is 'one'
    ),
    
    # Main panel for displaying outputs ----
    mainPanel(
      conditionalPanel(
        condition = "input.choice == 'one'",
        h3(textOutput("singleoutput")),
        p()
      ),
      
      conditionalPanel(
        condition = "input.choice != 'one'",
        # Output: Data file ----
        textOutput("message"), #%>% withSpinner(type=7, color="#957D6E", size = 0.5),
        textOutput("head_presentation"),
        #withSpinner(tableOutput("head"), type=7, color="#957D6E", size = 0.5),
        tableOutput("head"),
        p(),
        textOutput("description_presentation"),
        tableOutput("description"),
        p(),
        textOutput("CM_presentation"),
        conditionalPanel(
          condition = "input.choice == 'many'",
          p(),
          #h3("Classification summay"),
          tableOutput("classes_many"), p(),
          plotOutput("densiplot_many"),
          p(),p()
        ),
        conditionalPanel(
          condition = "input.choice == 'validation'",
          p(),
          plotOutput("CM"),
          plotOutput("densiplot"),
          plotOutput("caliplot"),
          p(),p()
        ),
        textOutput("down_presentation"),
        conditionalPanel(
          condition = "output.outputReady",
          downloadButton("downloadData", "Download the dataset...")
        ),
        p()#,
)
      # textOutput("CMstats_presentation"),
      # tableOutput("CMstats")
    )
    
  ) # closes sidebarLayout(
) # closes ui <- fluidPage(

# Define server logic to read selected file ----
server <- function(input, output, session) {
  
  get_DS <-reactive({
    if(is.null(input$dataset)) { return(NULL) }
    DS <- read.csv(file=input$dataset$datapath)
    return(DS)
  })
  
  output$fileUploaded <- reactive({
    return(!is.null(get_DS()))
  })  
  outputOptions(output, 'fileUploaded', suspendWhenHidden=FALSE)

  output$outputReady <- reactive({
    return(!is.null(exporter()))
  })  
  outputOptions(output, 'outputReady', suspendWhenHidden=FALSE)
  
  observeEvent(input$dataset, {
    DS <- get_DS()
    varnames <- colnames(DS)
    numvars <- c("", varnames)
    facvars <- c("", varnames)
    updateSelectInput(session, inputId = "input_Age_name", label="Age (in days)", choices = numvars)
    updateSelectInput(session, inputId = "input_HR_name", label="Heart rate (in beats/min)", choices = numvars)
    updateSelectInput(session, inputId = "input_MUAC_name", label="MUAC (in cm)", choices = numvars)
    updateSelectInput(session, inputId = "input_OxySat_name", label="Oxygen saturation (in %)", choices = numvars)
    updateSelectInput(session, inputId = "input_Lethargy_name", label="Lethargy", choices = facvars)
    updateSelectInput(session, inputId = "input_Death_name", label="Mortality", choices = facvars)
  })

  observeEvent(input$input_Lethargy_name, {
    if(input$input_Lethargy_name!="") {
      DS <- get_DS()
      chs <- c("", levels(as.factor(DS[,input$input_Lethargy_name])))
      updateSelectInput(session, inputId = "lethNoLabel", label="No", choices = chs)
      updateSelectInput(session, inputId = "lethYesLabel", label="Yes", choices = chs)
    }
  })

  observeEvent(input$input_Death_name, {
    if(input$input_Death_name!="") {
      DS <- get_DS()
      chs <- c("", levels(as.factor(DS[,input$input_Death_name])))
      updateSelectInput(session, inputId = "deathNoLabel", label="No", choices = chs)
      updateSelectInput(session, inputId = "deathYesLabel", label="Yes", choices = chs)
    }
  })
  
  
  output$message <-reactive({
    if(input$choice == "many") {
      validate(need(length(input$dataset$name)>0, "Please load your dataset."))
      validate(
        need(nchar(input$input_Age_name)>0 & nchar(input$input_HR_name)>0 & nchar(input$input_MUAC_name)>0 & 
               nchar(input$input_OxySat_name)>0 & nchar(input$input_Lethargy_name)>0, 
             paste("Please specify the column names identifying each variable"))
      )
      validate(
        need(length(unique(c(input$input_Age_name, input$input_HR_name, input$input_MUAC_name, 
                             input$input_OxySat_name, input$input_Lethargy_name)))==5, 
             paste("The same column name can not be indicated for two variables."))
      )
      validate(need(nchar(input$lethNoLabel)>0 & nchar(input$lethYesLabel)>0, "Please specify the labels that identify if there was lethargy or not."))
      validate(
        need(length(unique(c(input$lethNoLabel, input$lethYesLabel)))==2, 
             paste("The same label can not be indicated for both conditions (lethargy yes/no)."))
      )
      
    } else if(input$choice == "validation") {
      validate(need(length(input$dataset$name)>0, "Please load your dataset."))
      validate(
        need(nchar(input$input_Age_name)>0 & nchar(input$input_HR_name)>0 & nchar(input$input_MUAC_name)>0 & 
               nchar(input$input_OxySat_name)>0 & nchar(input$input_Lethargy_name)>0 & nchar(input$input_Death_name)>0, 
             paste("Please specify the column names identifying each variable"))
      )
      validate(
        need(length(unique(c(input$input_Age_name, input$input_HR_name, input$input_MUAC_name, 
                             input$input_OxySat_name, input$input_Lethargy_name, input$input_Death_name)))==6, 
             paste("The same column name can not be indicated for two variables."))
      )
      validate(need(nchar(input$lethNoLabel)>0 & nchar(input$lethYesLabel)>0, "Please specify the labels that identify if there was lethargy or not."))
      validate(
        need(length(unique(c(input$lethNoLabel, input$lethYesLabel)))==2, 
             paste("The same label can not be indicated for both conditions (lethargy yes/no)."))
      )
      validate(need(nchar(input$deathNoLabel)>0 & nchar(input$deathYesLabel)>0, "Please specify the labels that identify if the subject died or not."))
      validate(
        need(length(unique(c(input$deathNoLabel, input$deathYesLabel)))==2, 
             paste("The same label can not be indicated for both conditions (death yes/no)."))
      )
    }
    #return("passed")
  })
  
  silent_checker <- reactive({
    
      if (input$choice == 'many') {
        validate(
          need(length(input$dataset$name)>0, FALSE),
          need(nchar(input$input_Age_name)>0 & nchar(input$input_HR_name)>0 & nchar(input$input_MUAC_name)>0 & 
                 nchar(input$input_OxySat_name)>0 & nchar(input$input_Lethargy_name)>0, FALSE),
          need(length(unique(c(input$input_Age_name, input$input_HR_name, input$input_MUAC_name, 
                               input$input_OxySat_name, input$input_Lethargy_name)))==5, FALSE),
          need(nchar(input$lethNoLabel)>0 & nchar(input$lethYesLabel)>0, FALSE),
          need(length(unique(c(input$lethNoLabel, input$lethYesLabel)))==2, FALSE)
        )
      } else if (input$choice == 'validation') {
        validate(
          need(length(input$dataset$name)>0, FALSE),
          need(nchar(input$input_Age_name)>0 & nchar(input$input_HR_name)>0 & nchar(input$input_MUAC_name)>0 & 
                 nchar(input$input_OxySat_name)>0 & nchar(input$input_Lethargy_name)>0 & nchar(input$input_Death_name)>0, FALSE),
          need(length(unique(c(input$input_Age_name, input$input_HR_name, input$input_MUAC_name, 
                               input$input_OxySat_name, input$input_Lethargy_name, input$input_Death_name)))==6, FALSE),
          need(nchar(input$lethNoLabel)>0 & nchar(input$lethYesLabel)>0, FALSE),
          need(length(unique(c(input$lethNoLabel, input$lethYesLabel)))==2, FALSE),
          need(nchar(input$deathNoLabel)>0 & nchar(input$deathYesLabel)>0, FALSE),
          need(length(unique(c(input$deathNoLabel, input$deathYesLabel)))==2, FALSE)
        )
      }
    #return(TRUE)
  })
  
  
  get_WS <-reactive({
    silent_checker()
      
    req(input$dataset)
    df <- read.csv(input$dataset$datapath)#,
                   # header = input$header,
                   # sep = input$sep,
                   # quote = input$quote)
    Lethargy <- rep(NA, nrow(df))
    Lethargy[which(df[[input$input_Lethargy_name]] == input$lethNoLabel)] <- 0
    Lethargy[which(df[[input$input_Lethargy_name]] == input$lethYesLabel)] <- 1
    Lethargy <- as.factor(Lethargy)
    levels(Lethargy) <- c("No", "Yes")
    
    if(input$choice == 'validation') {
      dth <- rep(NA, nrow(df))
      dth[which(df[[input$input_Death_name]] == input$deathNoLabel)] <- 0
      dth[which(df[[input$input_Death_name]] == input$deathYesLabel)] <- 1
      dth <- as.factor(dth)
      levels(dth) <- c("No", "Yes")
      ws <- data.frame(Age = df[[input$input_Age_name]],
                       Heart.Rate = df[[input$input_HR_name]],
                       MUAC = df[[input$input_MUAC_name]],
                       Oxy.Sat = df[[input$input_OxySat_name]],
                       Lethargy = Lethargy,
                       Death = dth)
    } else if(input$choice == 'many') {
      ws <- data.frame(Age = df[[input$input_Age_name]],
                       Heart.Rate = df[[input$input_HR_name]],
                       MUAC = df[[input$input_MUAC_name]],
                       Oxy.Sat = df[[input$input_OxySat_name]],
                       Lethargy = Lethargy)
    }
    
    return(ws)
  })
  
  output$head_presentation <- renderText({
    if(input$choice == 'many' | input$choice == 'validation') {
      silent_checker()
      return("Dataset preview (first lines...):")
    } 
  })
  
  output$head <- renderTable({
    WS <- get_WS()
    return(head(WS))
  })
  
  output$description_presentation <- renderText({
    silent_checker()
    return("Dataset summary: mean (SD) and prevalences")
  })

  output$description <- renderTable({
    WS <- get_WS()
    if (input$choice == 'many') {
      leth.table <- table(WS$Lethargy, useNA="always")
      ct <- paste0(names(leth.table), ": ", leth.table, " (", round(leth.table/sum(leth.table), 1), "%)")
      varnames <- names(WS)
      means <- c(paste0(round(mean(WS$Age), 2), " (", round(sd(WS$Age), 2), ")"),
                 paste0(round(mean(WS$Heart.Rate), 2), " (", round(sd(WS$Heart.Rate), 2), ")"),
                 paste0(round(mean(WS$MUAC), 2), " (", round(sd(WS$MUAC), 2), ")"),
                 paste0(round(mean(WS$Oxy.Sat), 2), " (", round(sd(WS$Oxy.Sat), 2), ")"),
                 paste0(ct[1], "; ", ct[2]))
      nas <- c(sum(is.na(WS$Age)),
               sum(is.na(WS$Heart.Rate)),
               sum(is.na(WS$MUAC)),
               sum(is.na(WS$Oxy.Sat)),
               ct[3])
      description.table <- data.frame(Variable = varnames, Summary=means, Missings = nas)
    } else if (input$choice == 'validation') {
      leth.table <- table(WS$Lethargy, useNA="always")
      ct <- paste0(names(leth.table), ": ", leth.table, " (", round(leth.table/sum(leth.table), 1), "%)")
      death.table <- table(WS$Death, useNA="always")
      dt <- paste0(names(death.table), ": ", death.table, " (", round(death.table/sum(death.table), 1), "%)")
      varnames <- names(WS)
      means <- c(paste0(round(mean(WS$Age), 2), " (", round(sd(WS$Age), 2), ")"),
                 paste0(round(mean(WS$Heart.Rate), 2), " (", round(sd(WS$Heart.Rate), 2), ")"),
                 paste0(round(mean(WS$MUAC), 2), " (", round(sd(WS$MUAC), 2), ")"),
                 paste0(round(mean(WS$Oxy.Sat), 2), " (", round(sd(WS$Oxy.Sat), 2), ")"),
                 paste0(ct[1], "; ", ct[2]),
                 paste0(dt[1], "; ", dt[2]))
      nas <- c(sum(is.na(WS$Age)),
               sum(is.na(WS$Heart.Rate)),
               sum(is.na(WS$MUAC)),
               sum(is.na(WS$Oxy.Sat)),
               ct[3],
               dt[3])
      description.table <- data.frame(Variable = varnames, Summary=means, Missings = nas)
    }
    return(description.table)
  })
  
  get_WSclean <-reactive({
    WS <- get_WS()
    WSclean <- na.omit(as.data.table(WS))
    return(WSclean)
  })
    
  output$CM_presentation <- renderText({
    WS0 <- get_WS()
    WS  <- get_WSclean()
    msg <- paste0((nrow(WS0)-nrow(WS)), " rows omitted because they had missing values. The final dataset contains ", nrow(WS), " rows...")
    return(msg)
  })

  get_model <-reactive({
    if(is.null(input$dataset) & !input$input_submit) { return(NULL) }
    MRCmodel <- readRDS(file="MRCmodel2021.rds")
    return(MRCmodel)
  })

  make_pred <- reactive({
    WS <- get_WSclean()
    #caret.model <- get_model()
    model_data <- get_model() #readRDS(file="MRCmodel2021.rds")
    caret.model <- model_data$caret.model
    best_thresh <- model_data$best_thresh
    
    WSb <- WS; WSb$Lethargy <- as.numeric(WSb$Lethargy)-1
    predicted_p <- predict.train(caret.model, newdata = WSb, type="prob")
    predictions <- as.numeric(predicted_p[,2] > best_thresh$threshold)
    
    PS <- cbind(WS, Pred.prob=predicted_p[,2], Predictions = predictions)
  })  
  

  make_single_pred <- eventReactive(input$input_submit, {
    model_data <- get_model() #readRDS(file="MRCmodel2021.rds")
    caret.model <- model_data$caret.model
    best_thresh <- model_data$best_thresh
    dataline <- data.frame(Age = input$input_Age,
                           Heart.Rate = input$input_HR,
                           MUAC = input$input_MUAC,
                           Oxy.Sat = input$input_OxySat,
                           Lethargy = as.numeric(input$input_Lethargy))
    predicted_p <- predict.train(caret.model, newdata = dataline, type="prob")
    predictions <- as.numeric(predicted_p[2] > best_thresh$threshold)
    levels_predictions <- c("Low risk", "High risk")
    
    out <- paste("Predicted probability:", round(predicted_p[2], 3), "; Risk classication:", levels_predictions[predictions+1])
    return(out)
  })
  output$singleoutput <- renderText({
    return(make_single_pred())
  })
  

  output$CM <- renderPlot({
    if(input$choice == 'validation') {
      PS <- make_pred()
      PS$Predictions <- as.factor(PS$Predictions)
      levels(PS$Predictions) <- c("No", "Yes")
      cm <- (confusionMatrix(data=PS$Predictions, reference = PS$Death, positive = "Yes", mode="everything"))
      cmplot <- draw_confusion_matrix(cm)
      return(cmplot)
    }
  })

  output$classes_many <- renderTable({
    PS <- make_pred()
    # model_data <- get_model() 
    # best_thresh <- model_data$best_thresh
    class <- c("Low risk", "High risk")
    ns <- table(PS$Predictions)
    props <- paste0(round(ns/sum(ns)*100, 1), "%")
    classtable <- data.frame(class, freq=as.matrix(ns), props, row.names = NULL)
    return(classtable)
  })
    
  output$densiplot_many <- renderPlot({
    PS <- make_pred()
    model_data <- get_model() 
    best_thresh <- model_data$best_thresh
    densiplot <- ggplot(data = PS, aes(x = PS$Pred.prob)) + 
      geom_density(alpha = 0.2) +
      geom_vline(xintercept = best_thresh$threshold) +
      ylab("Density") + xlab(paste("predicted probability")) +
      ggtitle("Density plot") +
      #scale_fill_discrete(name = "Real outcome", labels = c("Alive", "Dead")) +
      theme_bw() + 
      theme(legend.position = "bottom") +
      annotate("label", x = best_thresh$threshold, y=3.35, label="threshold") +
      annotate("text" , x = best_thresh$threshold-0.01, y=3.0, label="Alive", hjust="right") +
      annotate("text" , x = best_thresh$threshold+0.01, y=3.0, label="Dead", hjust="left")
    return(densiplot)
  })
    
  output$densiplot <- renderPlot({
    PS <- make_pred()
    model_data <- get_model() 
    best_thresh <- model_data$best_thresh
    densiplot <- ggplot() + 
      geom_density(aes(x = PS$Pred.prob, fill = PS$Death), alpha = 0.2) +
      geom_vline(xintercept = best_thresh$threshold) +
      ylab("Density") + xlab(paste("predicted probability")) +
      ggtitle("Density plot") +
      #scale_fill_discrete(name = "Real outcome", labels = c("Alive", "Dead")) +
      theme_bw() + scale_fill_grey(name = "Real outcome", labels = c("Alive", "Dead")) + 
      theme(legend.position = "bottom") +
      annotate("label", x = best_thresh$threshold, y=3.35, label="threshold") +
      annotate("text" , x = best_thresh$threshold-0.01, y=3.0, label="Alive", hjust="right") +
      annotate("text" , x = best_thresh$threshold+0.01, y=3.0, label="Dead", hjust="left")
    return(densiplot)
  })

  
  output$caliplot <- renderPlot({
    if(input$choice == 'validation') {
      PS <- make_pred()
      model_data <- get_model() 
      best_thresh <- model_data$best_thresh
      cal_obj <- calibration(PS$Death ~ PS$Pred.prob, class="Yes", cuts=10)
      plot(cal_obj, type = "l", auto.key = list(lines = TRUE, points = FALSE))
      caliplot <- ggplot(cal_obj, type = "l", auto.key = list(lines = TRUE, points = FALSE)) +
        ggtitle("Calibration plot") +
        geom_vline(xintercept = best_thresh$threshold*100) +
        annotate("label", x = best_thresh$threshold*100, y=50, label="threshold") +
        annotate("text" , x = best_thresh$threshold*100-1, y=47.5, label="Alive", hjust="right") +
        annotate("text" , x = best_thresh$threshold*100+1, y=47.5, label="Dead", hjust="left") +
        theme_bw() +
        scale_x_continuous(breaks=seq(0,100,10)) +
        ylim(0,50)  +
        xlab(paste("predicted probability")) 
      return(caliplot)
    }
  })
  
  # output$CMstats <- renderTable({
  #   PS <- make_pred()
  #   PS$Predictions <- as.factor(PS$Predictions)
  #   levels(PS$Predictions) <- c("No", "Yes")
  #   cm <- (confusionMatrix(data=PS$Predictions, reference = PS$Death, positive = "Yes", mode="everything"))
  #   preout <- c(cm$overall, cm$byClass)
  #   out <- data.frame(Statistic = names(preout), Value = round(preout, 2))
  # })
    
  output$down_presentation <- renderText({
    silent_checker()
    return("You can download the sub-dataset with the predicted probabilities and outcomes. (Only works if running in browser, i.e. not the RStudio preview)")
  })

  exporter <- reactive({
    PS <- make_pred()
    PS$Predictions <- as.factor(PS$Predictions)
    levels(PS$Predictions) <- c("No", "Yes")
    return(PS)
  })

  output$downloadData <- downloadHandler(
    #print(str(exporter())),
    filename = function(){ 
      paste("MRC2020pred.csv") 
    },
    content = function(file) {
      write.csv(exporter(), file, row.names = FALSE)
    }
  )
  
  # output$raw <- renderTable({
  #   PS <- make_pred()
  #   PS$Predictions <- as.factor(PS$Predictions)
  #   levels(PS$Predictions) <- c("No", "Yes")
  #   return(PS[1:100,])
  # })
}
# Run the app ----
shinyApp(ui, server)
# runApp(shinyApp(ui, server), launch.browser = TRUE)
```
